# Model-based assessment of SARS-CoV-2 Delta variant transmission dynamics within partially vaccinated K-12 school populations

**DOI:** 10.1101/2021.08.20.21262389

**Authors:** Jennifer R. Head, Kristin L. Andrejko, Justin V. Remais

**Author notes:** Corresponding author: Justin V. Remais, PhD.

## Abstract

**Background:** We examined school reopening policies amidst rising transmission of the highly transmissible Delta variant, accounting for vaccination among individuals aged 12 years and older, with the goal of characterizing risk to students and teachers under various within-school non-pharmaceutical interventions (NPIs) combined with specific vaccination coverage levels.

**Methods:** We developed an individual-based transmission model to simulate transmission of the Delta variant of SARS-CoV-2 among a synthetic population, representative of Bay Area cities. We parameterized the model using community contact rates from vaccinated households ascertained from a household survey of Bay Area families with children conducted between February – April, 2021.

**Interventions and outcomes:** We evaluated the additional infections in students and teachers/staff resulting over a 128-day semester from in-school instruction compared to remote instruction when various NPIs (mask use, cohorts, and weekly testing of students/teachers) were implemented in schools, across various community-wide vaccination coverages (50%, 60%, 70%), and student (≥12 years) and teacher/staff vaccination coverages (50% - 95%). We quantified the added benefit of universal masking over masking among unvaccinated students and teachers, across varying levels of vaccine effectiveness (45%, 65%, 85%), and compared results between Delta and Alpha variant circulation.

**Results:** The Delta variant sharply increases the risk of within-school COVID-transmission when compared to the Alpha variant. In our highest risk scenario (50% community and within-school vaccine coverage, no within-school NPIs, and predominant circulation of the Delta variant), we estimated that an elementary school could see 33-65 additional symptomatic cases of COVID-19 over a four-month semester (depending on the relative susceptibility of children <10 years). In contrast, under the Bay Area reopening plan (universal mask use, community and school vaccination coverage of 70%), we estimated excess symptomatic infection attributable to school reopening among 2.0-9.7% of elementary students (8-36 excess symptomatic cases per school over the semester), 3.0% of middle school students (13 cases per school) and 0.4% of high school students (3 cases per school). Excess rates among teachers attributable to reopening were similar. Achievement of lower risk tolerances, such as <5 excess infections per 1,000 students or teachers, required a cohort approach in elementary and middle school populations. In the absence of NPIs, increasing the vaccination coverage of community members from 50% to 70% or elementary teachers from 70% to 95% reduced the estimated excess rate of infection among elementary school students attributable to school transmission by 24% and 41%, respectively. We estimated that with 70% coverage of the eligible community and school population with a vaccine that is ≤65% effective, universal masking can avert more cases than masking of unvaccinated persons alone.

**Conclusions:** Amidst circulation of the Delta variant, our findings demonstrated that schools are not inherently low risk, yet can be made so with high community vaccination coverages and universal masking. Vaccination of adult community members and teachers protects unvaccinated elementary and middle school children. Elementary and middle schools that can support additional interventions, such as cohorts and testing, should consider doing so, particularly if additional studies find that younger children are equally as susceptible as adults to the Delta variant of SARS-CoV-2.

**Limitations:** We did not consider the effect of social distancing in classrooms, or variation in testing frequency, and considerable uncertainty remains in key transmission parameters.

## Introduction

Assessments of the impacts of school closures and risks of reopening continue to be of high priority as school districts plan for the fall 2021 semester and more transmissible variants dominate the ongoing COVID-19 pandemic [1]. While school closures are intended to curb the spread of COVID-19, major risks for children’s mental health and educational and social development have been documented [2–4]. Introduction of vaccines with high effectiveness against infection [5–8] with SARS-CoV-2 reduce the risk of transmission within school environments in two ways. First, community transmission rates—which strongly impact the probability of within-school transmission [9]—are suppressed in areas with high vaccination coverage, although vaccination coverage remains heterogeneous across school districts [10]. Second, teachers, high school students, and some middle school students are now eligible for vaccination, conferring direct protection against within-school transmission. Prior epidemiological study and model-based risk assessments found the vaccine-eligible school population has higher risk of school-based transmission as compared to vaccine-ineligible elementary students [9, 11–13].

Nevertheless, due to rising rates of the more transmissible Delta variant across the U.S. [14], particularly in settings with low vaccination rates, there is concern that a return to schooling could be accompanied by increased risks of transmission [15], particularly among elementary school populations who are not yet eligible for vaccination. While our understanding of the natural history parameters for children is limited to the parent strain and thus evolving with continued Delta circulation, elementary-aged students (aged 5-10) may be less susceptible to infection than older children and adults [16–19]. Further, children under 18 have milder outcomes than adults [20], and exhibit extremely low fatality rates from SARS-CoV-2 (2 deaths per million by one estimate [21]) even among children with comorbidities [21]. However, there is already evidence that Delta’s enhanced infectivity is increasing rates of infection among US children, concurrent with the launch of the fall 2021 semester, especially in areas with low vaccination coverage and no mask mandates [22]. While most cases in children are mild, there are rare but serious cases of long-term sequelae that persist after COVID-19 infection in children, including Multisystem Inflammatory Syndrome (MIS) [23].

Guidance issued late summer 2021 from the Centers for Disease Prevention and Control (CDC) as well as the California Department of Public Health (CDPH) urged K-12 schools to fully reopen for instruction during the fall 2021 semester with masks required indoors for all students and staff [24, 25]. Spacing of at least three feet between students is also recommended, but if this cannot be achieved, it is recommended to apply layers of additional prevention measures, such as additional asymptomatic testing, symptom screening, or hand washing.

In March of 2020, the California Bay Area was among the first in the nation to close schools, moving the 2020 spring semester to remote instruction [26]. As of June 2021, California remained the state with the lowest percentage of students engaged in in-person instruction [27], and large Bay Area school districts, including San Francisco and Oakland, launched very limited in-person activities from April to June of 2021. Previous work has estimated the effect of initial closure for the 2020 spring semester on COVID-19 cases, hospitalizations and deaths in students, teachers, family members, and community members, and has examined the effect of reopening under various strategies on COVID-19 outcomes across a new four-month semester [9]. With the 2021 fall semester forthcoming, we examine questions surrounding school reopening in the context of an increasingly vaccinated population of individuals 12 years and older. We expand a previously published model [9] to include vaccination of adults in the community, teachers/staff, and students aged 12 years and older in order to examine which additional prevention measures beyond vaccination are required to limit excess cases to fewer than two student cases per school (<50% probability of a case per month) [28]. We also estimate whether achieving high levels of within-school vaccination coverages for teachers and students over 12 years would allow schools to safely drop additional prevention measures while maintaining low transmission. Finally, we quantify the additional benefit of universal masking compared to masking only among the unvaccinated, as a function of varied vaccine effectiveness. We examine scenarios assuming circulation of the highly transmissible Delta variant, and compare to outcomes estimated for the Alpha variant.

## Methods

We adapted a previously described agent-based model [9] to estimate the effect of fall 2021 reopening strategies under various vaccination coverages and SARS-CoV-2 variants, including the highly transmissible Delta variant. The model was informed by longitudinal data collected on children’s social contacts, including data on post-vaccination contact rates of children and their adult family members during spring 2021 school closures.

### Survey methodology and analysis

To parameterize community contact rates among the school-aged population and their adult family members within the model, we implemented a social contact survey of school-aged children in nine Bay Area counties (Alameda, Contra Costa, Marin, Napa, San Francisco, San Mateo, Santa Clara, Solano, Sonoma), as described elsewhere [9]. Survey respondents (one adult per household) reported the number and location of non-household contacts they and all of their children made within six age categories (0–4, 5–12, 13–17, 18–39, 40–64 and 65+ years) throughout the day prior. A contact was defined as an interaction within six feet lasting over five seconds [29]. Eligible households contained at least one school-aged child (pre-kindergarten to grade 12). We recruited participants using an online panel provider (Qualtrics) to be representative of Bay Area on the basis of race/ethnicity and income. The survey was implemented between February, 8 – April 1, 2021, when most (92%) of children were in remote schooling, and during a period where Bay Area healthcare workers, educators, and emergency personnel were eligible for COVID-19 vaccination.

### Transmission model

We generated 1,000 synthetic populations representative of the demographic composition of major Bay Area cities [30], in which we assigned each individual an age, household, and occupation status (student, teacher, school staff, other employment, not employed), as well as membership in a school or workplace. We separated schools into elementary (grades K–5; ages 5-10 years), middle (grades 6-8; ages 11-13) and high (grades 9-12; ages 14-17) schools, and assigned individuals grades and classrooms within each school, based on age. All individuals interacted with all other individuals in one of six ways, according to a hierarchy of highest shared membership: household > classroom or workplace > grade > school > community [31]. Age-specific community contact rates used in the simulation were obtained from surveys of households where at least one adult was vaccinated, as these individuals from these households had higher contact rates than individuals from unvaccinated households.

A discrete-time, age-structured, individual-based stochastic model was used to simulate SARS-CoV-2 transmission dynamics in the synthetic population (Figure 1A). At each time increment (one day), each individual was associated with an epidemiological state: fully vaccinated (V), susceptible (*S*), exposed (*E*), asymptomatic (*A*), symptomatic with non-severe illness (*C*), symptomatic with severe illness (*H*_1_, *D*_1_) resulting in eventual hospitalization before recovery (*H*_2_) or hospitalization before death (*D*_2_), recovery (*R*) or death (*M*). The susceptible compartment included individuals who received the vaccine, yet remained susceptible to infection. Transmission was implemented probabilistically for contacts between susceptible (S) and infectious individuals in the asymptomatic (A) or symptomatic and non-hospitalized states (C, H1, D1). Movement of individual *i* on day *t* from a susceptible to exposed class was determined by a Bernoulli random draw with probability of success given by the force of infection, *λ*_i,t_:

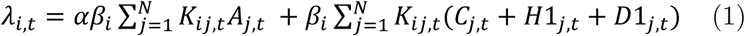

**Figure 1.**
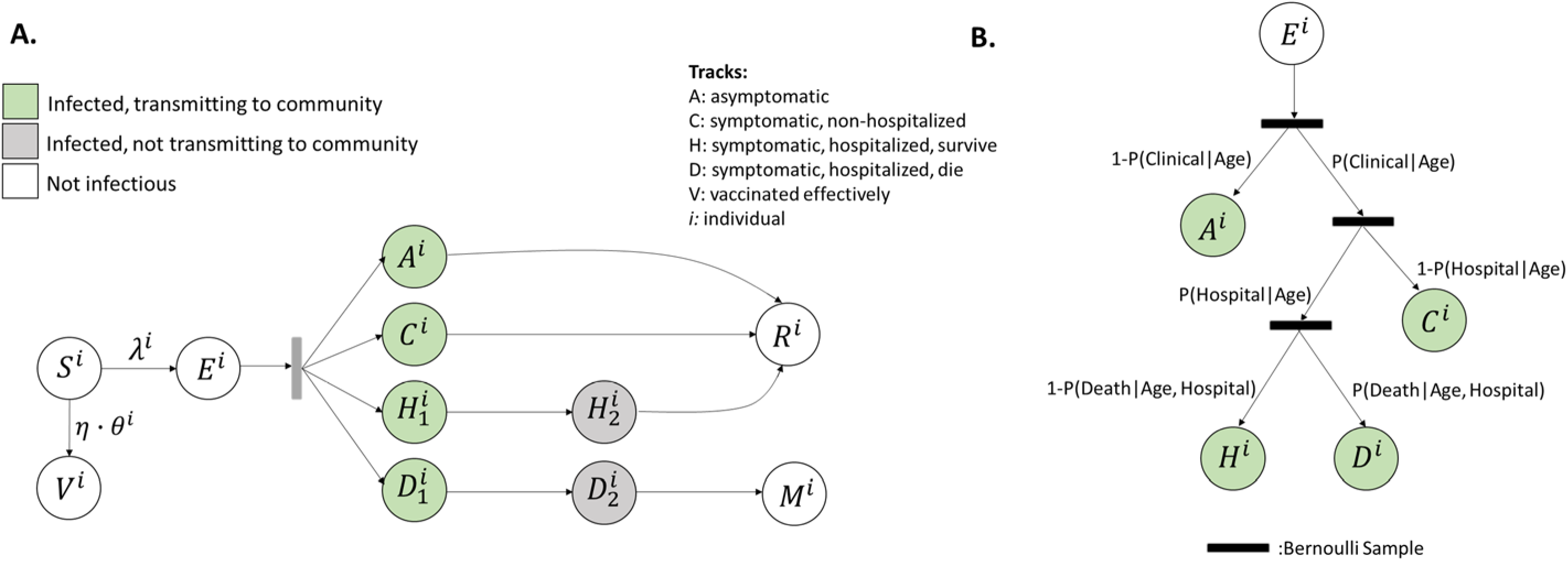
Model schematic (A) Schematic of the agent-based susceptible–exposed–infected–recovered (SEIR) model. S, susceptible; E, exposed; A, asymptomatic; C, symptomatic, will recover; H_1_, symptomatic and will recover, not yet hospitalized; H_2_, hospitalized and will recover; D_1_, symptomatic, not yet hospitalized; D_2_, hospitalized and will die; R, recovered; M, dead; V, vaccinated effectively; λ, force of infection defining movement from S to E; η, vaccine effectiveness, θ, vaccination coverage among subgroup to which individual *i* belongs. Superscript *i* refers to individual. After an agent enters the exposed class, they enter along their predetermined track, with waiting times between stage progression drawn from a Weibull distribution. V. (B) Schematic of the conditional probabilities by which agents are assigned a predetermined track.

where *N* is the number of individuals in the synthetic population (*N*=16,000), and *α* is the ratio of the transmissibility of asymptomatic individuals to symptomatic individuals. The fate followed by each individual after exposure was assigned from Bernoulli random draws at the start of each simulation based on age-stratified conditional probabilities (Figure 1B; Table S1). Once exposed, the duration of time spent in each disease stage were sampled from Weibull distributions (Table S1). Using estimates from studies evaluating risk of symptoms by age [18], we assumed 21% of infected individuals <20 years and 69% of infected individuals 20 years and older experienced symptoms [18]. Following previous work [18], we assumed *α* to be less than one, as asymptomatic individuals may be less likely to transmit infectious droplets by sneezing or coughing [32].

Based on an average of *R*_0_ for the Alpha (*R*_0_ = 2.5) and Delta (*R*_0_ = 5) variant weighted by the proportion of circulating variants in summer 2021 [33, 34], we calculated *R*_0_ as 4.6 and solved for the mean transmission rate of the pathogen, 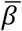, as the ratio between *R*_0_ and the product of the infection duration and the weighted mean number of daily contacts per individual during the pre-intervention period (Supporting Information, equation 2) [35]. To understand the influence of the Delta variant, we also ran simulations assuming full coverage of the Alpha variant. To represent age-varying susceptibility [18], we then calculated an age-stratified *β*_i_, that incorporated varying relative susceptibility by age while permitting the population mean to be 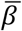 (Supporting Information, equations 3-4). We assumed that children under 10 years of age are half as susceptible to infection as older children and adults, in accordance with prior meta-analysis and modelling work reporting lower household secondary attack rates in children as compared to adults [16–19], with the lowest secondary attack rates in children less than 10 years of age [16, 19]. Nevertheless, given that some studies report equal susceptibility across all ages [36–38], and our current understanding of susceptibility is based largely on the Alpha variant, we also modeled scenarios without age-dependent susceptibility.

The daily contact rate between individuals *i* and *j* on day *t*, *K*_ij,t_, was estimated for pairs of individuals following previous study [31] based on their type of interaction (e.g., household, class, community). Contact rates were scaled by a time-dependent factor between 0 (complete closure) and 1 (no intervention) representing a social distancing intervention to reduce contact between individual pairs. Pairs with a school or workplace interaction were reassigned as community interactions under closures. Because symptomatic individuals mix less with the community [39], we incorporated isolation of symptomatic individuals and quarantine of their household members. Following prior work, we simulated a 100% reduction in daily school or work contacts and a 75% reduction in community contacts for a proportion of symptomatic individuals, and an additional proportion of their household members [40]. This means that a proportion of students and staff would stay home from school if they themselves were symptomatic, while a smaller percentage would stay home from school if one of their household members was symptomatic. We assumed that individuals were in the infectious class for up to three days prior to observing symptoms [41], during which time they did not reduce their daily contacts.

To establish the initial conditions for a new school semester, we simulated transmission continuously throughout three phases: 1) initiation of pandemic (schools open); 2) start of NPI enactment (schools closed for in-person instruction); 3) continuation of pandemic and NPIs across a long-term school closure period. This yielded 1,000 unique combinations of initial conditions. The simulated infection rates at the start of the semester ranged from 6-120 cases per 100,000, in accordance with infection rates among Bay Area counties in early August, 2021 [42]. The simulated infection seroprevalence at the start of the semester ranged from 1.5 to 10%, in line with seroprevalence data from San Francisco in late summer, 2020 [43]. Prior to simulating transmission over the school semester, a proportion of susceptible individuals aged 12 and older were moved to the vaccine compartment, according to a Bernoulli random draw with probability of success equal to the proportion vaccination coverage among the eligible population times the vaccine effectiveness. Vaccine effectiveness is set at 85% [44] for most simulations, but we also explore scenarios at lower effectiveness.

### Interventions and outcomes

#### Effect of within-school non-pharmaceutical interventions under various community vaccination coverages

We examined the effect of three non-pharmaceutical interventions across three levels of community vaccination coverage (50%, 60%, 70%), assuming that vaccination coverage within school children 12 years and older and teachers matches that in the community, and that the vaccine is 85% effective against infection [44]. First, we examined universal masking, assuming that the effectiveness of masks for reducing both inward and outward transmission [45] is 15% for elementary school students, 25% for middle school students, 35% for high school students, and 50% for teachers and staff [46–48]. Second, we examined a scenario of masking plus weekly testing of all students and teachers, in which we assumed a test with 85% sensitivity was administered every 7 days with 1 day to get results back [49]. We then assumed that the classroom and the household members of a positive test stayed home from school/work for 14 days and reduced community contacts by 75%. Third, we examined a masking plus cohort scenario in which classroom groups of 20 students were assumed to contact each other freely, with individuals within the cohort reducing their contacts with individuals outside their cohorts by 75%. While all of the nine Bay Area counties have achieved vaccination coverages of at least 60% as of summer, 2021, and some over 80% [50], we include the lower 50% to make the findings more generalizable to areas outside the Bay Area who may otherwise have similar demographics.

#### Effect of increasing vaccination among the school population in the absence of other interventions

Next, we considered within-school vaccination coverage in the absence of within-school NPIs (masking, testing, cohorting). We assumed a community vaccination coverage among the eligible population of 70%, which represented a conservative level of vaccination coverage among a Bay Area county [50]. We then examined COVID-19 outcomes if students 12 and older and teachers/staff had higher vaccination coverages (ranging from 70% to 95% coverage).

#### Effect of masking all individuals in a school compared to masking only unvaccinated individuals

Finally, we estimated the additional cases averted in each population by masking the entire student and teacher population, compared to masking only the unvaccinated student and teacher population, in the absence of additional interventions. We held community and within-school vaccination coverage of the eligible (12+) population at 70%, and varied vaccine effectiveness from 45% to 85%.

#### Outcomes

We evaluated two primary outcomes. Our first primary outcome was the increase in the total number of symptomatic infections among students and teachers/staff between in-school and remote instruction over a 128-day semester. We refer to this outcome as the excess symptomatic infections attributable to school transmission. We also examined the increase in the total number of all infections and hospitalization among students and teachers over the 128-day period between in-school and remote instruction.

Our second primary outcome was the minimum set of interventions required to maintain total excess infections among students and teachers attributable to school transmission below risk tolerances that may be relevant to decision-makers. We considered three school population-based risk tolerances, that varied in leniency from <5 to <50 additional cases per 1,000 school population. Following prior study, we examined a school-based risk tolerance of a monthly probability of an in-school transmission below 50% (<2 excess cases per school) [28].

We examined outcomes among population subgroups, focusing on students and teachers/staff, stratified by schooling level. We summarized all outcomes using the mean, median, and the 89^th^ percentile highest probability density interval (HPDI) across the 1,000 model realizations. 89% intervals are deemed to be more stable than the 95% intervals [51, 52].

### Ethics statement

Ethical approval was obtained from the Office for Protection of Human Subjects at the University of California, Berkeley (Protocol Number: 2020-04-13180). Prior to taking the anonymous survey, parents were provided details of the study and asked to provide written informed consent.

## Results

### Effect of within-school precautions under various community vaccination coverages (*children under 10 years half as susceptible to infection*)

We estimated higher rates of excess illness among elementary and middle school students as compared to high school students across all combinations of NPIs tested (Table 1; Table S6; Figure 2). Excess illness was also higher among elementary and middle school teachers, as compared to high school teachers, but differences between schooling levels were smaller among teachers as compared to students (Table S6; Figure 2). Increasing community and school vaccination coverage reduced excess illness attributable to school transmission among all populations, but particularly among the vaccine-eligible population (i.e., teachers and high school students) (Figure 2), both in the absence and presence of additional NPIs.

**Figure 2.**
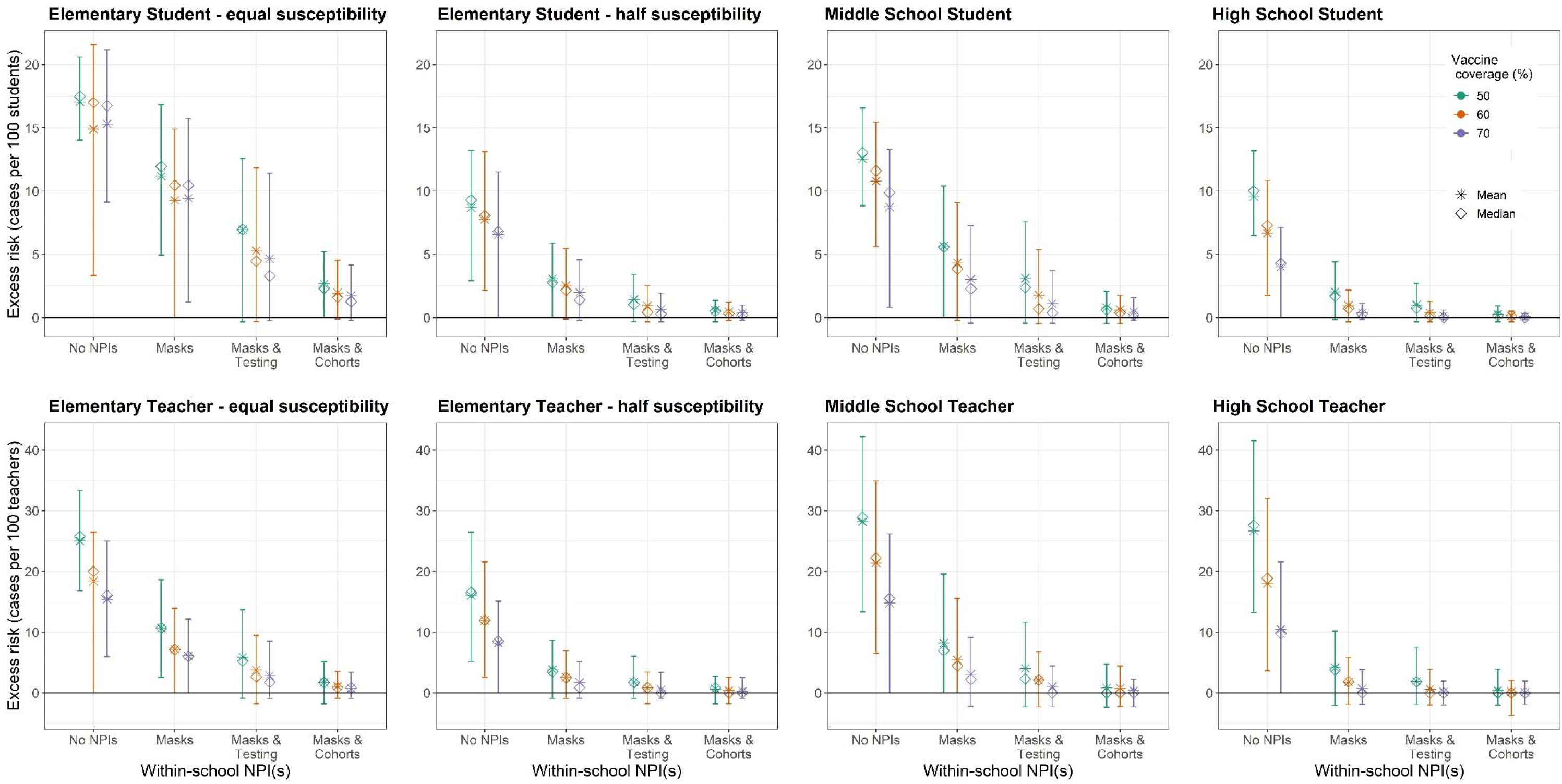
We examined the effect of three non-pharmaceutical interventions across three levels of community vaccination coverage (50%, 60%, 70%), assuming that vaccination coverage within school children 12+ and teachers matches that in the community and the vaccine effectiveness is 85% against symptomatic infection. Masks indicate universal masks regardless of vaccination status. We calculated the mean (stars) and median (diamonds) of excess cases per 100 persons attributable to school transmission among population subgroups across 1,000 model realizations. Vertical lines reflect the 89^th^percentile high probability density interval (HPDI).

**Table 1.** The number of excess student cases attributable to school transmission expected across a four-month (128-day) semester, for 70% community vaccination coverage, which is seen in most Bay Area counties [50]. The mask row is high<u>lighted to demonstrate the current minimum</u> required scenario for schools within the Bay Area.

|  | Excess student cases attributable to within-school transmission within: |  |  |  |
| --- | --- | --- | --- | --- |
|  | 380-person<br>elementary schools<br>(half susceptibility) | 380-person<br>elementary schools<br>(equal susceptibility) | 420-person<br>middle schools | 620-person high schools |
| No precautions | 25 cases per school | 57 cases per school | 37 cases per school | 27 cases per school |
| Universal masking | 8 cases per school | 36 cases per school | 13 cases per school | 3 cases per school |
| Masks + testing | 3 cases per school | 18 cases per school | 5 cases per school | 1 case per school |
| Masks + cohorts | 1 cases per school | 7 cases per school | 2 cases per school | 1 case per 2 schools |

Upon achieving a 70% community vaccination coverage or higher (the coverage observed in May 2021 in most Bay Area counties)[50] and without additional NPIs, we estimated the average excess incidence rate as between 4-9 symptomatic cases per 100 students across all age groups (Figure 2). Expressed as excess cases per school attributable to school transmission, this amounts to an estimated 27 excess cases per high school, 37 excess cases per middle school, and 25 excess symptomatic cases per elementary school across a 128-day semester (Table 1). Tables S2 and S3 display results for 50% and 60% vaccine coverage. Full results for symptomatic and asymptomatic infection are included in Tables S6 and S7.

Under the most likely reopening scenario for Bay Area schools – dominant circulation of the Delta variant, vaccination coverages of at least 70% and universal masks (Table 1; Figure 3) – we estimated an excess of eight symptomatic cases per elementary school, 13 cases per middle school, and three cases per high school attributable to school transmission over a 128-day semester. This equates to school-attributable illness in an additional 2.0% of elementary school students, 3.0% of middle school students, and 0.4% of high school students owing to school transmission. We estimated that an additional 1.7% of elementary school teachers, 3.1% of middle school teachers, and 0.7% of high school teachers would experience symptomatic infection attributable to school transmission across a semester.

**Figure 3.**
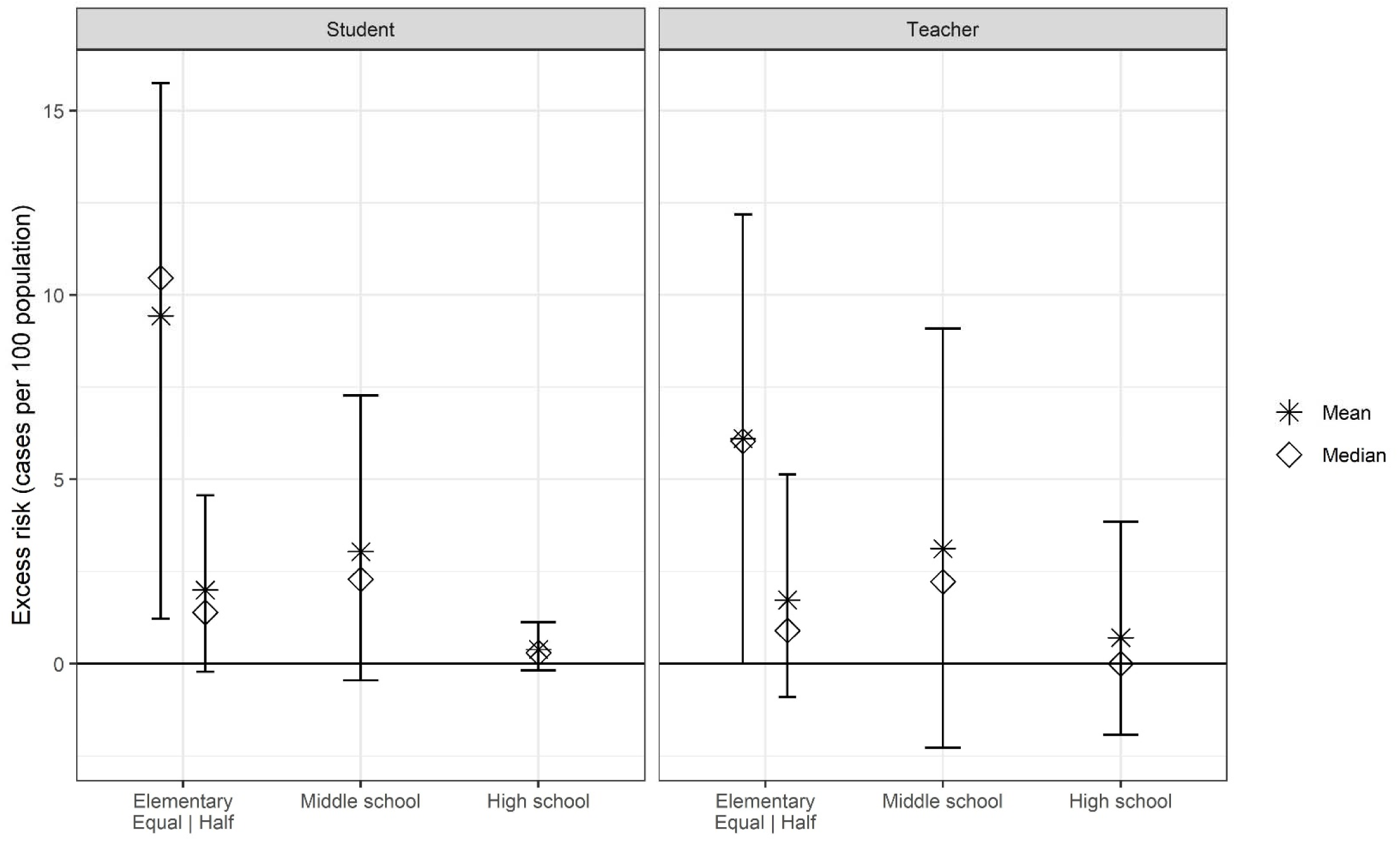
We projected the excess risk under the specific scenario likely to be observed during the Fall 2021 semester for the California Bay Area: **70% vaccination coverage among the eligible population and universal mask wearing** among all teachers and students. We assume 85% vaccine effectiveness against symptomatic infection. The mean (stars) and median(diamonds) of excess cases per 100 persons attributable to school transmission among population subgroups across 1,000 model realizations. Vertical lines reflect the 89^th^percentile high probability density interval (HPDI).

While children <12 years remain ineligible for vaccination, increasing vaccination among the community and teachers lowered risk of asymptomatic and symptomatic illness among young children. As simulated community vaccination coverage of the eligible population increased from 50% to 60% to 70%, we estimated that the estimated percent of elementary school children with a school-attributable symptomatic illness fell from 8.7% to 7.8% to 6.6%, representing a 24% decline in school-attributable transmission. This suggests that adult to child transmission represents an important source of school-attributable illnesses (Figure 2).

Within-school NPIs were most effective at reducing excess symptomatic cases within elementary and middle schools regardless of levels of community vaccination coverage, and within high schools with lower community vaccination coverages (Figure 2). For instance, where community vaccine coverage was 50% and no additional NPIs were taken, we estimated an excess incidence of 8.7 cases (89% HPDI: 2.9, 13.2) per 100 students in elementary schools, 12.5 (89% HPDI: 8.8, 16.6) per 100 students in middle schools and 9.6 per 100 students in high schools (89% HPDI: 6.5, 13.2). Adding masks but holding vaccine coverage constant, we estimated an excess incidence of 3.1 cases (89% HPDI: 0, 5.9) per 100 elementary students, 5.6 cases (89% HPDI: 0, 10.4) per 100 middle school students, and 2.0 (89% HPDI: -0.2, 4.4) cases per 100 high school students.

Across all model interventions scenarios and populations, hospitalizations and deaths were rare among students, but we simulated hospitalization in all scenarios except those that combined high vaccination (70%) with masks and cohorts. We simulated the highest estimated hospitalization rates among students of all grade levels when no NPIs were modelled. Following published estimates of age-dependent case hospitalization rate [53], our model assumes a higher probability of hospitalization among individuals aged 10 to 20 years compared to individuals under 10 years (Table S1). The maximum hospitalization rate simulated was 4.8 hospitalizations per one million middle school students over the 128-day semester, under 50% vaccination coverage and no additional precautions. The highest hospitalization rate simulated among elementary school students was 1.3 hospitalizations per one million elementary school students, under the same conditions, assuming elementary children are equally susceptible to infection as older children and adults. Simulated interventions combining masks and cohorts yielded hospitalization rates for the four-month semester under 3 per 10 million students, regardless of assumptions about susceptibility.

We estimated higher hospitalization rates among teachers and other school staff as compared to students. Under a 70% vaccine coverage scenario, excess hospitalizations among teachers was 48.6 per 100,000 teachers over the 128-day semester (daily rate: 0.40 per 100,000) without NPIs (Table 2). With the current universal mask recommendation, the excess hospitalization rate was 12.6 per 100,000 teachers per semester, or 0.09 per 100,000 per day. Adding a cohort approach to masking reduced the estimated excess hospitalization rate to 1.4 per 100,000 teachers per semester.

**Table 2.** The excess risk of hospitalization among all teachers (regardless of grade level) across a four-month school semester attributable to school transmission, depending on community vaccine coverage and modelling assumptions. Yellow row indicates the most likely scenario for the Bay Area fall 2021 reopening.

| NPIs | 50% coverage | 60% coverage | 70% coverage |
| --- | --- | --- | --- |
| **Hospitalization rate (per 100,000)** |  |  |  |
| None | 98.0 per 100,000 | 68.6 per 100,000 | 48.6 per 100,000 |
| Universal masking | 27.4 per 100,000 | 16.0 per 100,000 | 12.6 per 100,000 |
| Masks + testing | 13.9 per 100,000 | 7.0 per 100,000 | 4.8 per 100,000 |
| Masks + cohorts | 3.5 per 100,000 | 1.6 per 100,000 | 1.4 per 100,000 |

### Effect of increasing vaccination among the school population in the absence of other interventions

We examined under what vaccination coverages, if any, it might be possible to have a return to schooling without any additional NPIs (Figure 4). Increasing vaccination coverage of the eligible school population from 70% to 95% reduced mean estimates of excess cases among elementary students, suggesting that increasing vaccination coverage among elementary school teachers can reduce the force of infection among their students. For instance, increasing the vaccination coverage of the eligible school population (here, teachers) from 70% to 95% reduced the estimated excess rate of infection from 6.6 (89% HPDI: 0, 11.5) to 3.9 (89% HPDI: -0.2, 9.2) symptomatic cases per 100 elementary students across the four-month semester, representing a reduction of 41%. At the same time, increasing vaccination of teachers/staff from 70% to 95% reduced the estimated excess rate of infection among elementary teachers from 8.2 (89% HPDI: 0, 15.1) to 2.3 (89% HPDI: -0.9, 6.0) symptomatic cases per 100 teachers across the four-month semester, representing a reduction of 71%.

**Figure 4.**
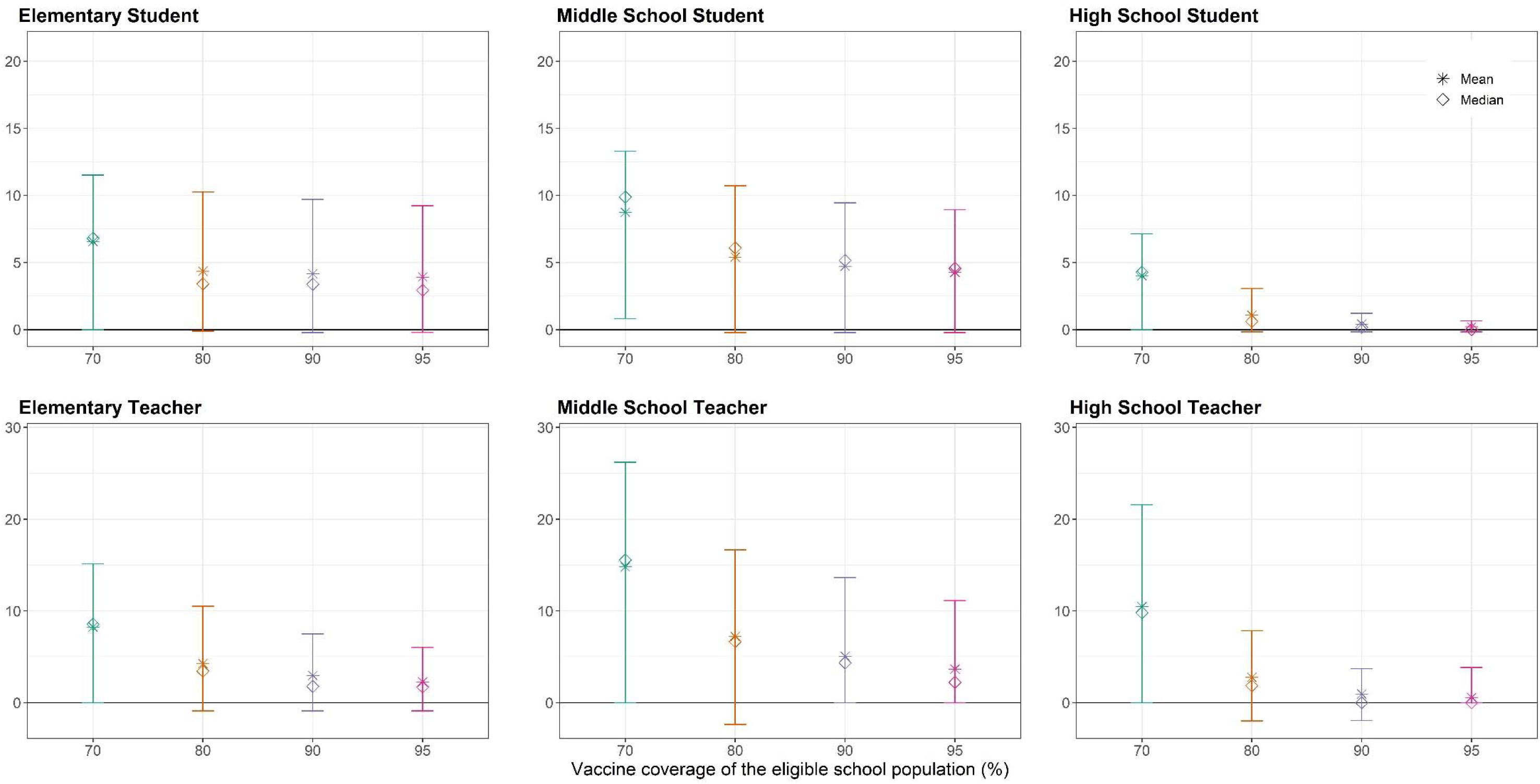
We examined the effect of increasing vaccination coverage among school populations, in the absence of additional non-pharmaceutical interventions, and holding community and within-school vaccination coverage of the eligible (12+) population at 70%. We calculated the mean (stars) and median(diamonds) of excess risk per 100 persons attributable to school transmission among population subgroups across 1,000 model realizations. Vertical lines reflect the 89^th^percentile high probability density interval (HPDI).

While increasing within-school vaccine coverage indirectly reduced infections among elementary and middle school students, the effect of increasing within-school vaccination coverage was most pronounced among high school students and teachers of all grade levels. Only high school teachers and students could achieve a transmission tolerance of fewer than 10 excess cases per 1,000 population attributable to school transmission using vaccination only without NPIs (Table 3). At 70% coverage of the eligible school population, we estimated an excess of 4.0 (89% HPDI: 0, 7.1) symptomatic cases per 100 students and 10.4 (89% HPDI: 0, 21.5) per 100 teachers across the 128-day semester, and at 95% coverage an excess of 0.2 (89% HPDI: -0.2, 0.6) cases per 100 students and 0.6 (89% HPDI: 0, 3.8) per 100 teachers across the 128-day semester (Figure 4). Among high school students and teachers/staff, we estimated a median of zero excess infections across the 1,000 model realizations when within-school coverages exceeded 90%.

**Table 3.** The minimum non-pharmaceutical intervention(s), or minimum within-school vaccination coverage of the eligible population, needed to reduce the risk of symptomatic infection to beneath a given risk level (e.g., 50 cases per 1,000 population), assuming that 70% of the vaccine-eligible community has received a vaccine at 85% effectiveness. ‘Not observed’ indicates that no combination of interventions *examined in this study* reduced excess risk beneath the indicated threshold. Masks refers to universal masking regardless of vaccination status.

|  |  | Population-wide risk tolerance — symptomatic cases per 1,000 population |  |  | School-based risk tolerance — < 2 cases per school* |
| --- | --- | --- | --- | --- | --- |
|  |  | <50 | <10 | <5 |  |
| Students | Elementary school – *half susceptibility* | Masks or 80% within-school coverage | Masks + testing | Masks + cohorts | Masks + cohorts |
|  | Elementary school – *equal susceptibility* | Masks + testing | Not observed** | Not observed** | Not observed** |
|  | Middle school | Masks or 90% within-school coverage | Masks + cohorts | Masks + cohorts | Masks + cohorts |
|  | High school | 70% within-school coverage | Masks or 90% within-school coverage | Masks or 90% within-school coverage | Masks + testing |
| Teachers/staff | Elementary school – *half susceptibility* | Masks or 80% within-school coverage | Masks + testing | Masks + cohorts |  |
|  | Elementary school – *equal susceptibility* | Masks + testing | Masks + cohorts | Not observed** |  |
|  | Middle school | Masks or 95% within-school coverage | Masks + cohorts | Masks + cohorts |  |
|  | High school | Masks or 80% within-school coverage | Masks or 90% within-school coverage | Masks + testing |  |

\*Assuming a 380-person elementary school, 420-person middle school, and 680-person high school
\*\*Not observed under the interventions examined here

### Interventions required to reduce incidence attributable within schools below certain risk tolerances

We examined whether layering NPIs or increasing within-school vaccination could reduce incidence attributable to school transmission below specific risk tolerances (Table 3). We estimated that universal masking and 70% community and within-school vaccination coverage or higher could reduce the number of excess cases attributable to school transmission to <50 per 1,000 students and teachers across all grade levels. In high school students, increasing the vaccine coverage among the vaccine-eligible school population above 70% could also reduce excess transmission to <50 per 1,000 students and teachers in the absence of NPIs. However, achieving lower risk levels among elementary school students—e.g., <10 cases per 1,000 students or teachers—required additional NPIs, such as testing or cohorts, and was not achievable through the NPIs investigated here if children under 10 years are equally as susceptible as adults. On a per school basis, reducing the excess cases attributable to school transmission to fewer than two cases per school across the full semester (i.e., <50% probability of a case per school per month) required both masks and cohorts. Among high schools, achieving this risk tolerance required combining masks with testing. Tables S2 and S3 display the minimum NPIs required to achieve the various risk tolerances assuming 50% and 60% vaccine coverage in the eligible community, respectively.

### Effect of masking all individuals in a school compared to masking only unvaccinated individuals

We compared the differences in school-attributable transmission under scenarios where only unvaccinated individuals wore masks compared to if all individuals masked, across different levels of vaccine effectiveness (VE), assuming 70% of the eligible population is fully vaccinated (Figure 5). Since all elementary students are unvaccinated, such a rule would change behaviors only among the vaccinated teachers, about 5% of the overall school population. In contrast, such a rule would affect the entirety of the vaccinated high school population, both students and teachers, about 70% of the overall school population. The difference between masking the entire student and teacher population as compared to only the unvaccinated school population is thus most apparent in middle and high school populations, and at lower VEs. For instance, given 45% VE, masking all middle and high school students and teachers would avert symptomatic infection for 3.9% of middle school students, 6.1% of high school students, 12.5% of middle school teachers, and 18.5% of high school teachers compared to masking only unvaccinated students and teachers. At 85% VE and above, there was little difference in school-attributable transmission between masking unvaccinated persons versus masking all persons.

**Figure 5.**
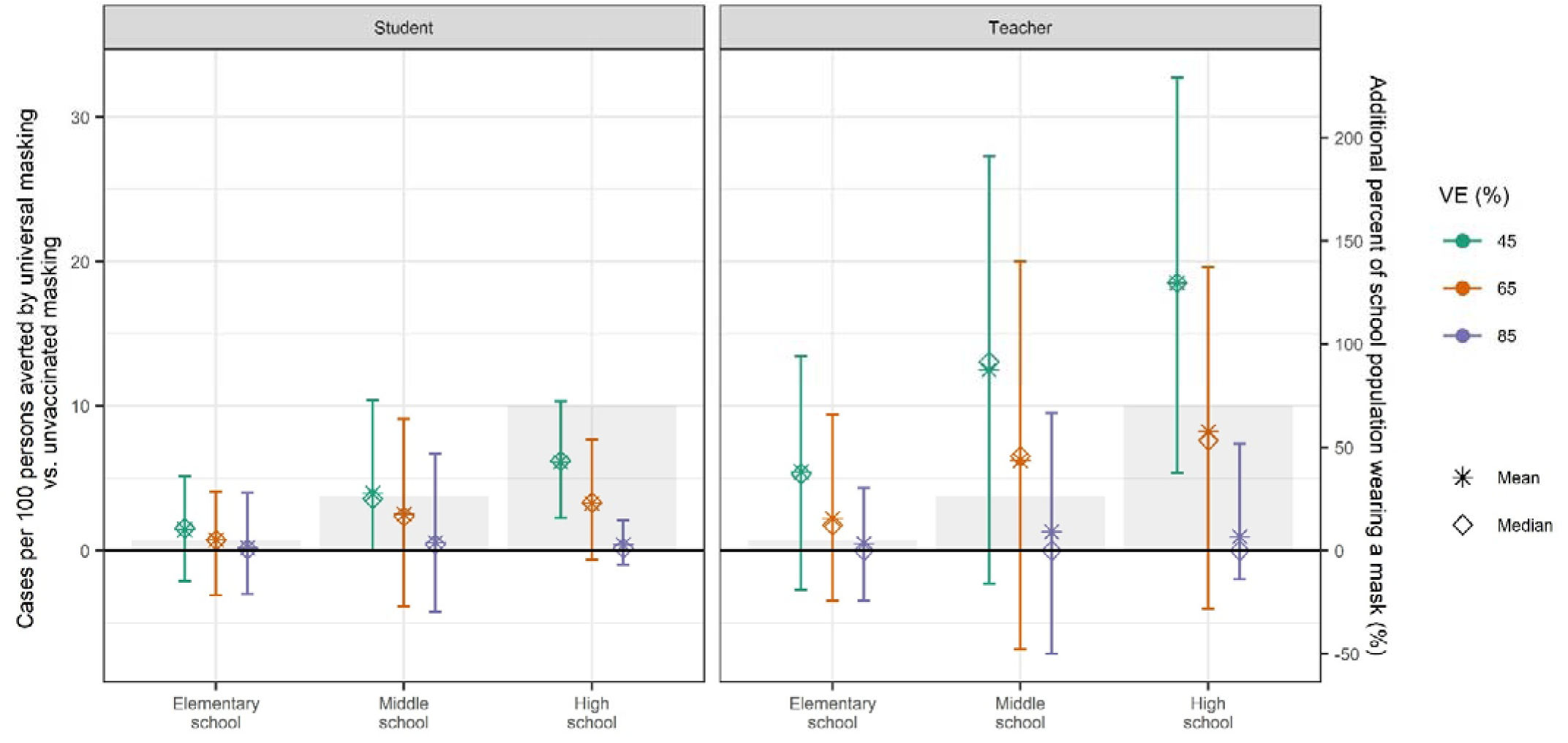
We estimated the additional cases averted in each population by masking the entire student and teacher population, compared to masking only the unvaccinated student and teacher population, in the absence of additional interventions. We held community and within-school vaccination coverage of the eligible (12+) population at 70%, and varied vaccine efficacy (VE). We calculated the mean (stars) and median(diamonds) of excess risk per 100 persons attributable to school transmission among population subgroups across 1,000 model realizations. Vertical lines reflect the 89^th^percentile high probability density interval (HPDI). Shaded bars and right axis reflect the vaccinated percent of the school population, for whom a universal masking rule as compared to a masking rule among the unvaccinated would apply.

### Key uncertainties

#### Susceptibility of children

We found that the excess risk in elementary schools is substantially altered if children under 10 years of age are considered equally as susceptible to SARS-CoV-2 as older children and adults when compared with half as susceptible (Figures 2 and 3; Tables 1 and 3). Under the current Bay Area reopening scenario (70% coverage + masks), the estimated number of within-school infections due to school transmission jumps from 8 to 36 cases per 380-person elementary school over the four-month semester under equal susceptibility assumptions. This corresponds to excess illness attributable to schools among 9.7% (89% HPDI: 1.2, 15.7%) of elementary students and among 6.1% (89% HPDI: 0, 1.2%) of elementary teachers. Only the most lenient risk tolerance of <50 excess infections per 1,000 elementary students (5%) was achievable with the combination of interventions examined here. The strictest combination of interventions tested (masks + cohorts, 70% vaccine coverage), would result in excess infection among 1.7% (89% HPDI: -0.2, 4.2) of elementary students, and 0.8% (89% HPDI: -0.9, 3.4) of elementary teachers.

The relative susceptibility of younger children to infection remains under debate, and the natural history parameters for emerging variants is evolving. Should younger children be as susceptible to SARS-CoV-2 as older children and adults, masking alone may not be sufficient to achieve low rates of transmission among elementary school populations.

#### Circulating variants of concern

Our results were highly sensitive to the proportion of variants of concern circulating. We examined outcomes if the Alpha variant had remained the dominant variant (*R*_0_ = 2.5), finding school attributable excess transmission to be nearly ten times lower than under circulation of the Delta variant when examining the most likely reopening scenario for this fall (70% vaccine coverage and universal masks) (Figure S1; Table S8). Under this scenario, we estimated fewer than one additional infection per school (<25% probability of an in-school transmission per month). At the level of community vaccination coverage observed in the Bay Area (70% coverage or higher), the most lenient risk tolerance of <50 additional cases per 1,000 students, was achievable without additional NPIs (Table S8; Figure S1). Under this no-NPI scenario, risk to the student population was estimated at 1 excess case per high school, 4 excess cases per middle school, and 1-5 excess cases (depending on susceptibility to SARS-CoV-2) per elementary school. We estimated that high schools could achieve very strict risk tolerances (<1 excess cases in 1,000 students) without any additional NPIs as long as vaccination coverage among the eligible school population exceeded 75% (Table S9).

We also projected fewer hospitalizations and deaths if the Alpha variant had remained the dominant variant. Under full circulation of the Alpha variant, we did not observe hospitalizations among students and observed very few hospitalizations among teachers within our model realizations. Under a 70% vaccine coverage scenario, excess hospitalizations among teachers was 23 per 100,000 (daily rate: 0.19 per 100,000) without any NPIs. When any school interventions were present (e.g., masking) under a 70% vaccine coverage scenario, our model realizations observed fewer than 1 excess teacher hospitalization per 100,000 teachers attributable to school transmission over the semester.

## Discussion

We simulated transmission of the Delta variant of SARS-CoV-2 in schools over an upcoming school semester with variable vaccine coverages within the school and community populations to approximate conditions that may be observed in the fall of 2021. Aligning with current CDPH and CDC reopening guidelines [24], which urge a full return to in-person schools with vaccination and universal mask usage, we estimated that an additional 0.4 to 3% of students, depending on schooling level, would experience symptomatic illness attributable to schools across the four month semester, with similar rates estimated for teachers (Figure 3). Under these scenarios, we estimated a daily school-attributable hospitalization rate as 0.09 per 100,000 teachers per day (Table 2). Vaccination is recognized by the CDC and CDPH as the leading public health strategy for reducing within-school transmission [24, 25], and our results highlight that increased vaccination coverage—both among the general community and among the eligible school population—plays an essential role in limiting symptomatic illness attributable to school transmission.

Our findings support the use of universal masks as precaution within schools, particularly elementary and middle schools, but also high schools that have within-school vaccine coverage <90%. Masks are supported as one of the simplest, yet effective, mitigation strategies [24, 25]. Masking is of particular importance for elementary and many middle school students who remain ineligible for vaccination; we estimated that a typical 380-person elementary school and 420-person middle school could see 25 and 37 symptomatic cases, respectively, of COVID-19 over the four-month semester under a reopening plan that did not involve masking (or other NPIs) and where community vaccination coverage is 70%. Using masks, even those that are only 15-25% effective, reduced that risk in our simulations to eight cases per elementary school and 13 per middle school per semester.

Nevertheless, achieving lower risk tolerances, such as fewer than ten additional school-attributable infections per 1,000 school population, required adding additional layers of protection, e.g., reduced contact between students via cohorting. This suggests that schools may want to consider additional precautions above and beyond the minimum requirement of masks. For instance, schools that can implement a cohort approach, or provide regular testing, should consider doing so. We estimated that high school students were at lower risk of infection, assuming vaccination rates among students matched those of the surrounding community, but nevertheless may require both masking and weekly testing to achieve a transmission probability of <50% per school per month.

Uncertainty was greatest among middle school and high school teachers and students, in part because pockets of low vaccine coverage within these environments can be sufficient to support occasional outbreaks. In our model, vaccine coverage was distributed randomly throughout the full population, such that some school realizations had vaccination coverage of teachers or students well below 50%, where transmission was possible. This represents reality, where certain school populations may have lower vaccination coverages than others.

Increased vaccine coverage of community members and teachers helped reduce illness among children not yet age-eligible for vaccination. We estimated that increasing vaccination coverage of the general population reduced the excess risk of transmission by 24% among elementary students. Similarly, we estimated that increasing vaccination coverage among teachers from 70% to 95% reduced the excess risk of school transmission by 41% among elementary students. This suggests that teacher to student transmission is an important route of transmission that can be eliminated by increased vaccination.

This study has limitations. First, community contact rates measured in the survey may reflect underestimates of actual community contact rates, as vaccination prevalence has increased since February – April of 2021 and community contacts increased with vaccination prevalence (survey data). Thus, our estimates of excess cases attributable to school transmission may be slight underestimates. Nevertheless, our previous work has shown that the effect of community transmission is minimized when within-school precautions are implemented [9]. Second, our transmission model does not capture all the precautions outlined in CDC guidance, including the effect of spacing desks three feet apart, or handwashing. Such precautions are difficult to model using a contact-based transmission model, and thus our estimates may overestimate risk if schools continue to emphasize such measures. Moreover, testing is an important component of the CDPH plan for return to in-person schooling. While we including testing as a potential NPI, we do not thoroughly investigate various testing routines that yield the optimal benefit, as does other work [54]. Third, most of our model simulations assume a high vaccine effectiveness against symptomatic COVID-19. Vaccine effectiveness may change as novel variants emerge and circulate; however, early studies indicate that vaccine effectiveness against variants of concern— including Delta—generally remain high [55]. Fourth, our initial conditions for seroprevalence in the synthetic population encompassed published 2020 seroprevelance estimates for the Bay Area, so may represent an underestimate of the true seroprevalence in the community as of 2021. If so, our modelled estimates of excess cases may represent a slight overestimate. Finally, our modelling results are sensitive to assumptions about the values for certain parameters, such as relative susceptibility of children to SARS-CoV-2, for which there remains high uncertainty. As the Delta variant poses unseen challenges to school communities, we do not have empirical data to support our model results. However, our estimates of the school-attributable risk are consistent with that reported by another modelling study [28], and consistent with increasing rates of infection among children, particularly in parts of the country with low vaccination coverage and no mask mandates in schools [22].

## Conclusion

Our findings support recommendations made by CDC and CDPH to fully reopen K-12 schools for the Fall 2021 semester; encourage high levels of vaccine uptake among eligible students and staff; and maintain mask usage, particularly among the unvaccinated elementary and middle school populations and high school populations with vaccine coverage <90%. Vaccination remains the most effective and sustainable means of risk reduction and efforts should focus on increasing vaccination coverage among the eligible community members and school population. Among populations not yet eligible for vaccination and communities with lower vaccination coverage, prevention measures, such as masking, may be needed to reduce the risk of school outbreaks. Schools may consider layering testing or cohorting as additional safety measures, particularly as the Delta variant takes hold.

## Data Availability

Data and code for this analysis can be found at https://github.com/jrhead/COVIDandVaccinatedSchools

## Acknowledgments

This research was supported in part by National Science Foundation grant no. 2032210, by National Institutes of Health grant nos. R01AI125842 and R01AI148336, and by the MIDAS Coordination Center (MIDASSUP2020-4) by a grant from the National Institute of General Medical Science (3U24GM132013-02S2).

## Supplemental text

**Table S1.**
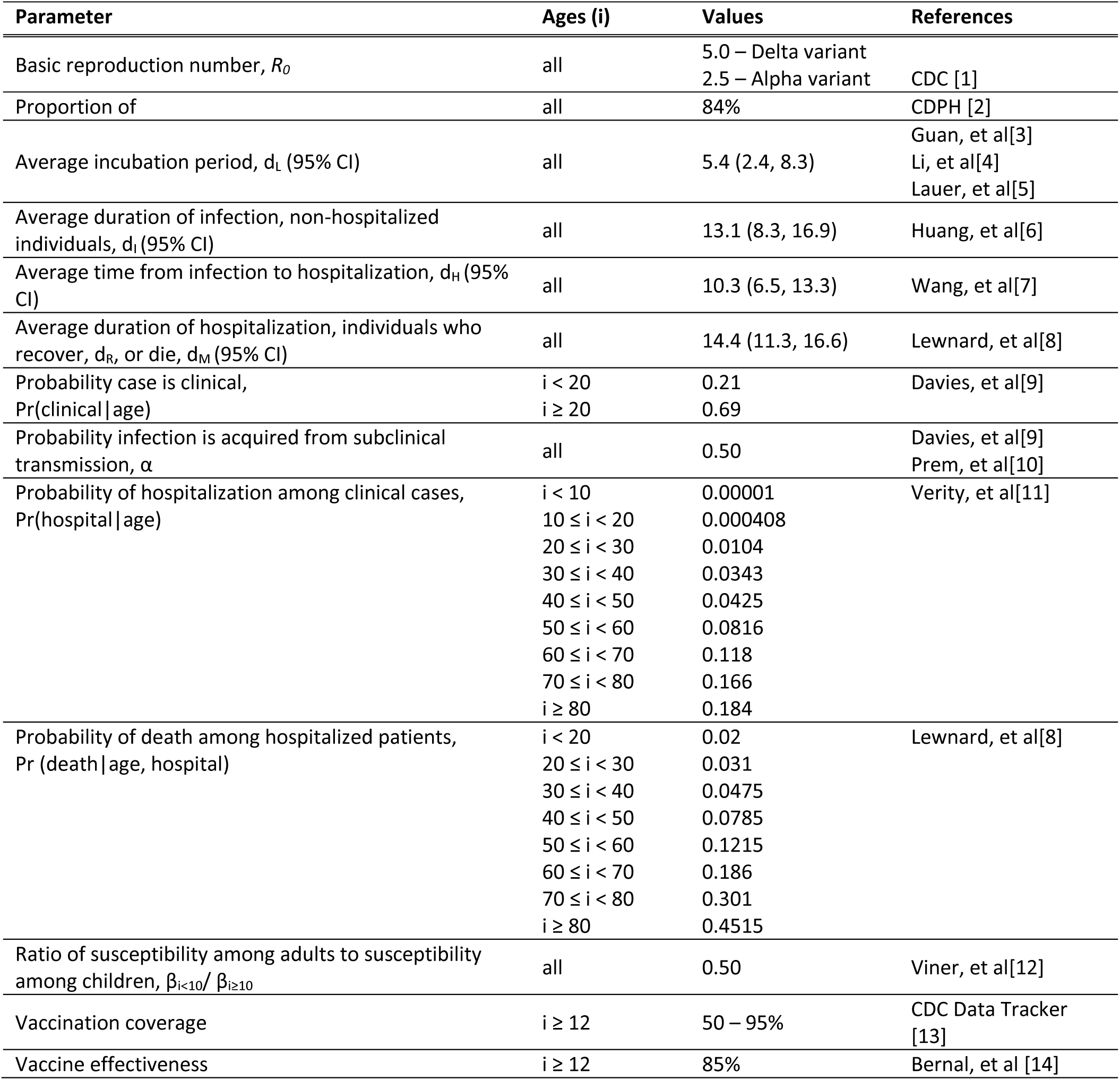
Parameters of the susceptible-exposed-infected-recovered model

| Parameter | Ages (i) | Values | References |
| --- | --- | --- | --- |
| Basic reproduction number, $R_0$ | all | 5.0 – Delta variant<br>2.5 – Alpha variant | CDC [1] |
| Proportion of | all | 84% | CDPH [2] |
| Average incubation period, $d_L$ (95% CI) | all | 5.4 (2.4, 8.3) | Guan, et al[3]<br>Li, et al[4]<br>Lauer, et al[5] |
| Average duration of infection, non-hospitalized individuals, $d_i$ (95% CI) | all | 13.1 (8.3, 16.9) | Huang, et al[6] |
| Average time from infection to hospitalization, $d_H$ (95% CI) | all | 10.3 (6.5, 13.3) | Wang, et al[7] |
| Average duration of hospitalization, individuals who recover, $d_R$, or die, $d_M$ (95% CI) | all | 14.4 (11.3, 16.6) | Lewnard, et al[8] |
| Probability case is clinical, $\Pr(\text{clinical} \vert \text{age})$ | $i < 20$<br>$i \geq 20$ | 0.21<br>0.69 | Davies, et al[9] |
| Probability infection is acquired from subclinical transmission, $\alpha$ | all | 0.50 | Davies, et al[9]<br>Prem, et al[10] |
| Probability of hospitalization among clinical cases, $\Pr(\text{hospital} \vert \text{age})$ | $i < 10$<br>$10 \leq i < 20$<br>$20 \leq i < 30$<br>$30 \leq i < 40$<br>$40 \leq i < 50$<br>$50 \leq i < 60$<br>$60 \leq i < 70$<br>$70 \leq i < 80$<br>$i \geq 80$ | 0.00001<br>0.000408<br>0.0104<br>0.0343<br>0.0425<br>0.0816<br>0.118<br>0.166<br>0.184 | Verity, et al[11] |
| Probability of death among hospitalized patients, $\Pr(\text{death} \vert \text{age, hospital})$ | $i < 20$<br>$20 \leq i < 30$<br>$30 \leq i < 40$<br>$40 \leq i < 50$<br>$50 \leq i < 60$<br>$60 \leq i < 70$<br>$70 \leq i < 80$<br>$i \geq 80$ | 0.02<br>0.031<br>0.0475<br>0.0785<br>0.1215<br>0.186<br>0.301<br>0.4515 | Lewnard, et al[8] |
| Ratio of susceptibility among adults to susceptibility among children, $\beta_{i < 10} / \beta_{i \geq 10}$ | all | 0.50 | Viner, et al[12] |
| Vaccination coverage | $i \geq 12$ | 50 – 95% | CDC Data Tracker [13] |
| Vaccine effectiveness | $i \geq 12$ | 85% | Bernal, et al [14] |

**Table S2.** The number of excess student cases attributable to school transmission expected across a four-month semester, for 50% community vaccination coverage.

|  | Excess student cases attributable to within-school transmission within: |  |  |  |
| --- | --- | --- | --- | --- |
|  | 380-person<br>**elementary schools**<br>(*equal susceptibility*) | 380-person<br>**elementary schools**<br>(*half susceptibility*) | 420-person **middle schools** | 620-person **high schools** |
| No precautions | 69 cases per school | 33 cases per school | 53 cases per school | 65 cases per school |
| Masks | 42 cases per school | 12 cases per school | 24 cases per school | 14 cases per school |
| Masks + testing | 26 cases per school | 5 cases per school | 13 cases per school | 7 cases per school |
| Masks + cohorts | 10 cases per school | 2 cases per school | 3 cases per school | 2 cases per school |

**Table S3.** The number of excess student cases attributable to school transmission expected across a four-month semester, for 60% community vaccination coverage.

|  | Excess student cases attributable to within-school transmission within: |  |  |  |
| --- | --- | --- | --- | --- |
|  | 380-person<br>**elementary schools**<br>(*equal susceptibility*) | 380-person<br>**elementary schools**<br>(*half susceptibility*) | 420-person **middle schools** | 620-person **high schools** |
| No precautions | 57 cases per school | 29 cases per school | 45 cases per school | 45 cases per school |
| Masks | 35 cases per school | 10 cases per school | 18 cases per school | 7 cases per school |
| Masks + testing | 20 cases per school | 4 cases per school | 8 cases per school | 3 cases per school |
| Masks + cohorts | 7 cases per school | 2 cases per school | 3 cases per school | 1 cases per school |

**Table S4.** The minimum non-pharmaceutical intervention needed to reduce the risk of symptomatic infection to beneath a given threshold (e.g., 50 cases per 1,000 population), assuming that 50% of the vaccine-eligible community has received a vaccine at 85% effectiveness. ‘Not observed’ indicates that no combination of interventions examined in this study reduced excess risk beneath the indicated threshold.

|  |  | Threshold - symptomatic cases per 1,000 population |  |  | < 2 cases per school* |
| --- | --- | --- | --- | --- | --- |
|  |  | <50 | <10 | <5 |  |
| Students | Elementary school – *half susceptibility* | Masks | Masks + cohorts | Not observed** | Not observed** |
|  | Elementary school – *equal susceptibility* | Masks + cohorts | Not observed** | Not observed** | Not observed** |
|  | Middle school | Masks + testing | Masks + cohorts | Not observed** | Not observed** |
|  | High school | Masks | Masks + cohorts | Masks + cohorts | Not observed** |
| Teachers | Elementary school – *half susceptibility* | Masks | Masks + cohorts | Not observed** |  |
|  | Elementary school – *equal susceptibility* | Masks + cohorts | Not observed** | Not observed** |  |
|  | Middle school | Masks + testing | Masks + cohorts | Not observed** |  |
|  | High school | Masks | Masks + cohorts | Masks + cohorts |  |

\*Assuming a 380-person elementary school, 420-person middle school, and 680-person high school
\*\*not observed under the specific combination of interventions simulated

**Table S5.** The minimum non-pharmaceutical intervention to reduce the risk of symptomatic infection to beneath a given threshold (e.g., 50 cases per 1,000 population), assuming that 60% of the vaccine-eligible community has received a vaccine at 85% effectiveness. ‘Not observed’ indicates that no combination of interventions examined in this study reduced excess risk beneath the indicated threshold.

|  |  | Threshold - symptomatic cases per 1,000 population |  |  | < 2 cases per school* |
| --- | --- | --- | --- | --- | --- |
|  |  | <50 | <10 | <5 |  |
| Students | Elementary school – *half susceptibility* | Masks | Masks + testing | Masks + cohorts | Masks + cohorts |
|  | Elementary school – *equal susceptibility* | Masks + cohorts | Not observed** | Not observed** | Not observed** |
|  | Middle school | Masks | Masks + cohorts | Not observed** | Not observed** |
|  | High school | Masks | Masks | Masks + testing | Masks + cohorts |
| Teachers | Elementary school – *half susceptibility* | Masks | Masks + testing | Masks + cohorts |  |
|  | Elementary school – *equal susceptibility* | Masks + testing | Masks + cohorts | Not observed** |  |
|  | Middle school | Masks + testing | Masks + cohorts | Not observed** |  |
|  | High school | Masks | Masks + testing | Masks + cohorts |  |

\*Assuming a 380-person elementary school, 420-person middle school, and 680-person high school
\*\*not observed under the specific combination of interventions simulated

**Table S6.** Excess *symptomatic* infections attributable to school transmission by population subgroup and scenario examined

| Vaccine coverage (%) | NPI | Susceptibility of children <10 years | Population | Mean (89% HPDI) excess infections per 100 population | Median excess infections per 100 population | Infections per school* |
| --- | --- | --- | --- | --- | --- | --- |
| 50 | None | Equal | Elementary student | 17.1 (14, 20.6) | 17.5 | 65 |
| 50 | None | Half | Elementary student | 8.7 (2.9, 13.2) | 9.3 | 33 |
| 50 | None |  | Middle school student | 12.5 (8.8, 16.6) | 13 | 53 |
| 50 | None |  | High school student | 9.6 (6.5, 13.2) | 10 | 65 |
| 50 | None | Equal | Elementary teacher | 25 (16.8, 33.3) | 25.8 |  |
| 50 | None | Half | Elementary teacher | 16.1 (5.2, 26.5) | 16.5 |  |
| 50 | None |  | Middle school teacher | 28.2 (13.3, 42.2) | 28.9 |  |
| 50 | None |  | High school teacher | 26.7 (13.2, 41.5) | 27.6 |  |
| 50 | Masks | Equal | Elementary student | 11.2 (4.9, 16.8) | 11.9 | 42 |
| 50 | Masks | Half | Elementary student | 3.1 (0, 5.9) | 2.8 | 12 |
| 50 | Masks |  | Middle school student | 5.6 (0, 10.4) | 5.6 | 24 |
| 50 | Masks |  | High school student | 2 (-0.2, 4.4) | 1.7 | 14 |
| 50 | Masks | Equal | Elementary teacher | 10.7 (2.6, 18.6) | 10.7 |  |
| 50 | Masks | Half | Elementary teacher | 3.9 (-0.9, 8.7) | 3.5 |  |
| 50 | Masks |  | Middle school teacher | 8.2 (0, 19.6) | 7 |  |
| 50 | Masks |  | High school teacher | 4.2 (-2.1, 10.2) | 3.8 |  |
| 50 | Masks + Testing | Equal | Elementary student | 6.9 (-0.3, 12.6) | 7 | 26 |
| 50 | Masks + Testing | Half | Elementary student | 1.4 (-0.3, 3.4) | 1 | 5 |
| 50 | Masks + Testing |  | Middle school student | 3.1 (-0.5, 7.6) | 2.4 | 13 |
| 50 | Masks + Testing |  | High school student | 1 (-0.3, 2.7) | 0.7 | 7 |
| 50 | Masks + Testing | Equal | Elementary teacher | 5.9 (-0.9, 13.7) | 5.3 |  |
| 50 | Masks + Testing | Half | Elementary teacher | 1.8 (-0.9, 6) | 1.7 |  |
| 50 | Masks + Testing |  | Middle school teacher | 4.1 (-2.3, 11.6) | 2.3 |  |
| 50 | Masks + Testing |  | High school teacher | 1.9 (-2, 7.5) | 1.9 |  |
| 50 | Masks + Cohorts | Equal | Elementary student | 2.7 (0, 5.2) | 2.3 | 10 |
| 50 | Masks + Cohorts | Half | Elementary student | 0.6 (-0.3, 1.4) | 0.5 | 2 |
| 50 | Masks + Cohorts |  | Middle school student | 0.8 (-0.5, 2.1) | 0.6 | 3 |
| 50 | Masks + Cohorts |  | High school student | 0.3 (-0.3, 0.9) | 0.2 | 2 |
| 50 | Masks + Cohorts | Equal | Elementary teacher | 1.7 (-1.8, 5.1) | 1.7 |  |
| 50 | Masks + Cohorts | Half | Elementary teacher | 0.7 (-1.8, 2.7) | 0.9 |  |
| 50 | Masks + Cohorts |  | Middle school teacher | 0.8 (-2.4, 4.8) | 0 |  |
| 50 | Masks + Cohorts |  | High school teacher | 0.4 (-2, 3.9) | 0 |  |
| 60 | None | Equal | Elementary student | 14.9 (3.3, 21.6) | 17 | 57 |
| 60 | None | Half | Elementary student | 7.8 (2.2, 13.1) | 8.1 | 29 |
| 60 | None |  | Middle school student | 10.8 (5.6, 15.5) | 11.6 | 45 |
| 60 | None |  | High school student | 6.7 (1.8, 10.8) | 7.3 | 45 |
| 60 | None | Equal | Elementary teacher | 18.4 (0, 26.5) | 20 |  |
| 60 | None | Half | Elementary teacher | 11.9 (2.6, 21.6) | 12 |  |
| 60 | None |  | Middle school teacher | 21.4 (6.5, 34.9) | 22.2 |  |
| 60 | None |  | High school teacher | 18 (3.6, 32.1) | 18.9 |  |
| 60 | Masks | Equal | Elementary student | 9.3 (0, 14.9) | 10.5 | 35 |
| 60 | Masks | Half | Elementary student | 2.6 (-0.1, 5.5) | 2.2 | 10 |
| 60 | Masks |  | Middle school student | 4.3 (-0.2, 9.1) | 3.8 | 18 |
| 60 | Masks |  | High school student | 1 (-0.3, 2.2) | 0.7 | 7 |
| 60 | Masks | Equal | Elementary teacher | 7.2 (0, 13.9) | 7.1 |  |
| 60 | Masks | Half | Elementary teacher | 2.6 (-0.9, 7) | 2.5 |  |
| 60 | Masks |  | Middle school teacher | 5.4 (0, 15.6) | 4.4 |  |
| 60 | Masks |  | High school teacher | 1.8 (-2, 5.9) | 1.9 |  |
| 60 | Masks + Testing | Equal | Elementary student | 5.3 (-0.3, 11.8) | 4.5 | 20 |
| 60 | Masks + Testing | Half | Elementary student | 0.9 (-0.3, 2.5) | 0.4 | 4 |
| 60 | Masks + Testing |  | Middle school student | 1.8 (-0.5, 5.4) | 0.7 | 8 |
| 60 | Masks + Testing |  | High school student | 0.4 (-0.3, 1.3) | 0.2 | 3 |
| 60 | Masks + Testing | Equal | Elementary teacher | 3.8 (-1.8, 9.5) | 2.7 | 14 |
| 60 | Masks + Testing | Half | Elementary teacher | 0.9 (-1.8, 3.4) | 0.9 | 3 |
| 60 | Masks + Testing |  | Middle school teacher | 2.1 (-2.3, 6.8) | 2.1 | 9 |
| 60 | Masks + Testing |  | High school teacher | 0.7 (-2, 3.9) | 0 | 5 |
| 60 | Masks + Cohorts | Equal | Elementary student | 2 (-0.1, 4.5) | 1.6 | 7 |
| 60 | Masks + Cohorts | Half | Elementary student | 0.5 (-0.2, 1.2) | 0.3 | 2 |
| 60 | Masks + Cohorts |  | Middle school student | 0.6 (-0.5, 1.8) | 0.4 | 3 |
| 60 | Masks + Cohorts |  | High school student | 0.2 (-0.3, 0.5) | 0.1 | 1 |
| 60 | Masks + Cohorts | Equal | Elementary teacher | 1.1 (-0.9, 3.6) | 0.9 |  |
| 60 | Masks + Cohorts | Half | Elementary teacher | 0.4 (-1.8, 2.6) | 0 |  |
| 60 | Masks + Cohorts |  | Middle school teacher | 0.8 (-2.3, 4.4) | 0 |  |
| 60 | Masks + Cohorts |  | High school teacher | 0.2 (-3.7, 2) | 0 |  |
| 70 | None | Equal | Elementary student | 15.3 (9.1, 21.2) | 16.8 | 58 |
| 70 | None | Half | Elementary student | 6.6 (0, 11.5) | 6.8 | 25 |
| 70 | None |  | Middle school student | 8.7 (0.8, 13.3) | 9.9 | 37 |
| 70 | None |  | High school student | 4 (0, 7.1) | 4.3 | 27 |
| 70 | None | Equal | Elementary teacher | 15.4 (6, 25) | 16.1 |  |
| 70 | None | Half | Elementary teacher | 8.2 (0, 15.1) | 8.5 |  |
| 70 | None |  | Middle school teacher | 16 (4.3, 28.9) | 16.3 |  |
| 70 | None |  | High school teacher | 10.1 (0, 20.4) | 9.6 |  |
| 70 | Masks | Equal | Elementary student | 9.4 (1.2, 15.7) | 10.5 | 36 |
| 70 | Masks | Half | Elementary student | 2 (-0.2, 4.6) | 1.4 | 8 |
| 70 | Masks |  | Middle school student | 3 (-0.4, 7.3) | 2.3 | 13 |
| 70 | Masks |  | High school student | 0.4 (-0.2, 1.1) | 0.3 | 3 |
| 70 | Masks | Equal | Elementary teacher | 6.1 (0, 12.2) | 6 |  |
| 70 | Masks | Half | Elementary teacher | 1.7 (-0.9, 5.1) | 0.9 |  |
| 70 | Masks |  | Middle school teacher | 3.1 (-2.3, 9.1) | 2.2 |  |
| 70 | Masks |  | High school teacher | 0.7 (-1.9, 3.8) | 0 |  |
| 70 | Masks + Testing | Equal | Elementary student | 4.7 (-0.2, 11.4) | 3.3 | 18 |
| 70 | Masks + Testing | Half | Elementary student | 0.7 (-0.3, 1.9) | 0.3 | 3 |
| 70 | Masks + Testing |  | Middle school student | 1.1 (-0.4, 3.7) | 0.4 | 5 |
| 70 | Masks + Testing |  | High school student | 0.1 (-0.2, 0.6) | 0 | 1 |
| 70 | Masks + Testing | Equal | Elementary teacher | 2.9 (-0.9, 8.5) | 1.8 |  |
| 70 | Masks + Testing | Half | Elementary teacher | 0.5 (-0.9, 3.4) | 0 |  |
| 70 | Masks + Testing |  | Middle school teacher | 1 (-2.3, 4.4) | 0 |  |
| 70 | Masks + Testing |  | High school teacher | 0.2 (-2, 2) | 0 |  |
| 70 | Masks + Cohorts | Equal | Elementary student | 1.7 (-0.2, 4.2) | 1.3 | 7 |
| 70 | Masks + Cohorts | Half | Elementary student | 0.4 (-0.2, 1) | 0.2 | 1 |
| 70 | Masks + Cohorts |  | Middle school student | 0.5 (-0.2, 1.6) | 0.2 | 2 |
| 70 | Masks + Cohorts |  | High school student | 0.1 (-0.2, 0.3) | 0 | 1 |
| 70 | Masks + Cohorts | Equal | Elementary teacher | 0.8 (-0.9, 3.4) | 0.9 |  |
| 70 | Masks + Cohorts | Half | Elementary teacher | 0.3 (-0.9, 2.5) | 0 |  |
| 70 | Masks + Cohorts |  | Middle school teacher | 0.4 (-2.3, 2.3) | 0 |  |
| 70 | Masks + Cohorts |  | High school teacher | 0.1 (-2, 2) | 0 |  |

\*Assuming a 380-person elementary school, 420-person middle school, and 680-person high school

**Table S7.**
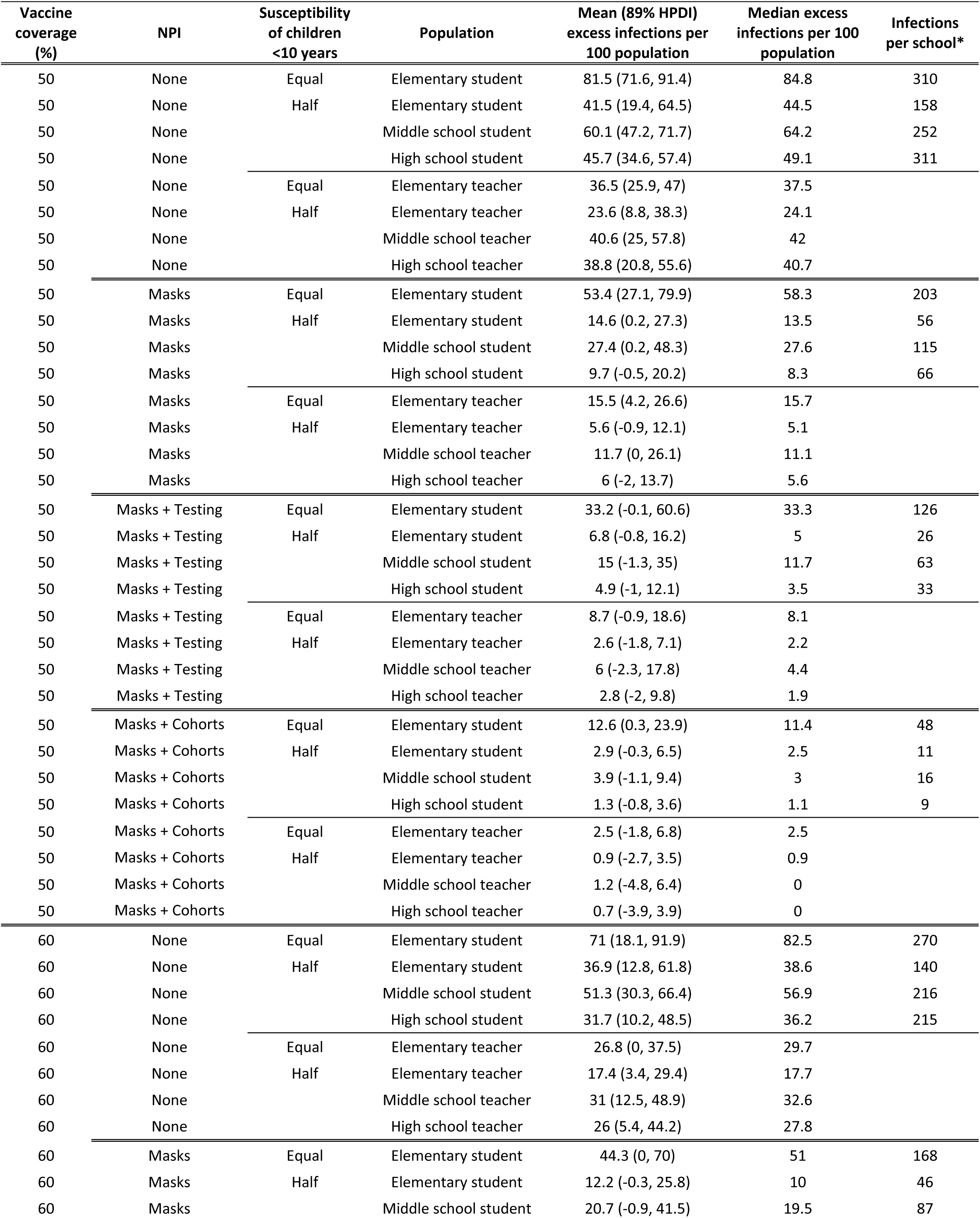
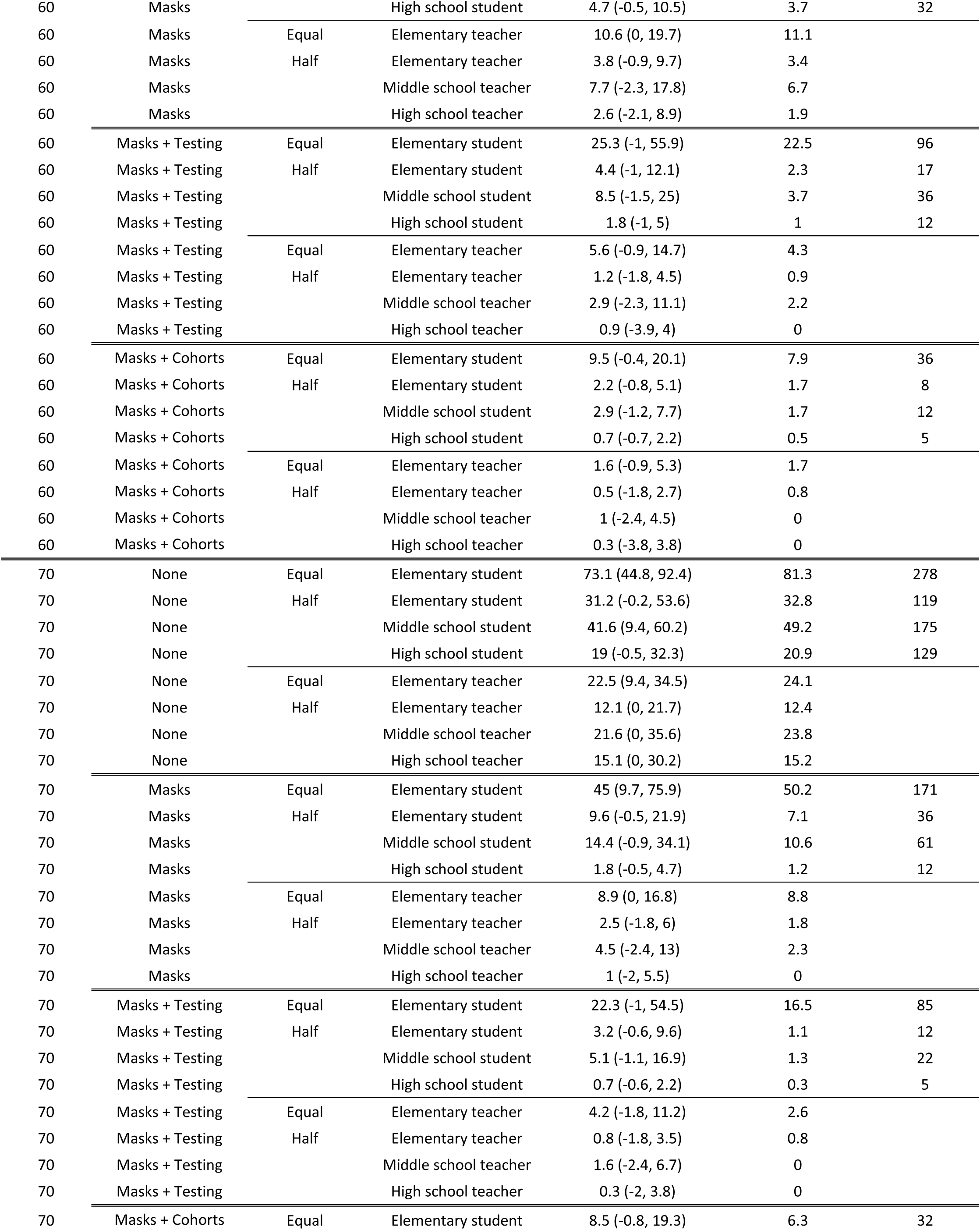
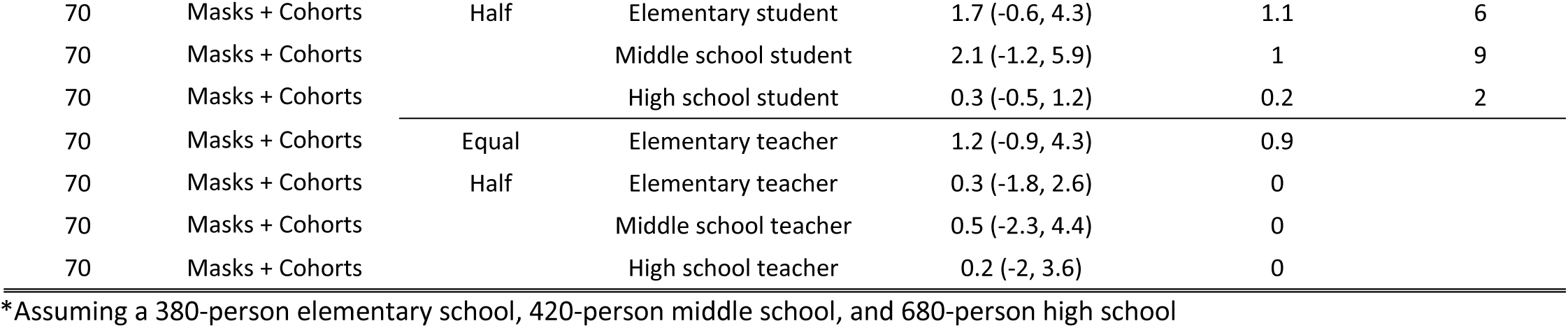
Excess infections (including asymptomatic infection) attributable to school transmission by population subgroup and scenario examined

| Vaccine coverage (%) | NPI | Susceptibility of children <10 years | Population | Mean (89% HPDI) excess infections per 100 population | Median excess infections per 100 population | Infections per school* |
| --- | --- | --- | --- | --- | --- | --- |
| 50 | None | Equal | Elementary student | 81.5 (71.6, 91.4) | 84.8 | 310 |
| 50 | None | Half | Elementary student | 41.5 (19.4, 64.5) | 44.5 | 158 |
| 50 | None |  | Middle school student | 60.1 (47.2, 71.7) | 64.2 | 252 |
| 50 | None |  | High school student | 45.7 (34.6, 57.4) | 49.1 | 311 |
| 50 | None | Equal | Elementary teacher | 36.5 (25.9, 47) | 37.5 |  |
| 50 | None | Half | Elementary teacher | 23.6 (8.8, 38.3) | 24.1 |  |
| 50 | None |  | Middle school teacher | 40.6 (25, 57.8) | 42 |  |
| 50 | None |  | High school teacher | 38.8 (20.8, 55.6) | 40.7 |  |
| 50 | Masks | Equal | Elementary student | 53.4 (27.1, 79.9) | 58.3 | 203 |
| 50 | Masks | Half | Elementary student | 14.6 (0.2, 27.3) | 13.5 | 56 |
| 50 | Masks |  | Middle school student | 27.4 (0.2, 48.3) | 27.6 | 115 |
| 50 | Masks |  | High school student | 9.7 (-0.5, 20.2) | 8.3 | 66 |
| 50 | Masks | Equal | Elementary teacher | 15.5 (4.2, 26.6) | 15.7 |  |
| 50 | Masks | Half | Elementary teacher | 5.6 (-0.9, 12.1) | 5.1 |  |
| 50 | Masks |  | Middle school teacher | 11.7 (0, 26.1) | 11.1 |  |
| 50 | Masks |  | High school teacher | 6 (-2, 13.7) | 5.6 |  |
| 50 | Masks + Testing | Equal | Elementary student | 33.2 (-0.1, 60.6) | 33.3 | 126 |
| 50 | Masks + Testing | Half | Elementary student | 6.8 (-0.8, 16.2) | 5 | 26 |
| 50 | Masks + Testing |  | Middle school student | 15 (-1.3, 35) | 11.7 | 63 |
| 50 | Masks + Testing |  | High school student | 4.9 (-1, 12.1) | 3.5 | 33 |
| 50 | Masks + Testing | Equal | Elementary teacher | 8.7 (-0.9, 18.6) | 8.1 |  |
| 50 | Masks + Testing | Half | Elementary teacher | 2.6 (-1.8, 7.1) | 2.2 |  |
| 50 | Masks + Testing |  | Middle school teacher | 6 (-2.3, 17.8) | 4.4 |  |
| 50 | Masks + Testing |  | High school teacher | 2.8 (-2, 9.8) | 1.9 |  |
| 50 | Masks + Cohorts | Equal | Elementary student | 12.6 (0.3, 23.9) | 11.4 | 48 |
| 50 | Masks + Cohorts | Half | Elementary student | 2.9 (-0.3, 6.5) | 2.5 | 11 |
| 50 | Masks + Cohorts |  | Middle school student | 3.9 (-1.1, 9.4) | 3 | 16 |
| 50 | Masks + Cohorts |  | High school student | 1.3 (-0.8, 3.6) | 1.1 | 9 |
| 50 | Masks + Cohorts | Equal | Elementary teacher | 2.5 (-1.8, 6.8) | 2.5 |  |
| 50 | Masks + Cohorts | Half | Elementary teacher | 0.9 (-2.7, 3.5) | 0.9 |  |
| 50 | Masks + Cohorts |  | Middle school teacher | 1.2 (-4.8, 6.4) | 0 |  |
| 50 | Masks + Cohorts |  | High school teacher | 0.7 (-3.9, 3.9) | 0 |  |
| 60 | None | Equal | Elementary student | 71 (18.1, 91.9) | 82.5 | 270 |
| 60 | None | Half | Elementary student | 36.9 (12.8, 61.8) | 38.6 | 140 |
| 60 | None |  | Middle school student | 51.3 (30.3, 66.4) | 56.9 | 216 |
| 60 | None |  | High school student | 31.7 (10.2, 48.5) | 36.2 | 215 |
| 60 | None | Equal | Elementary teacher | 26.8 (0, 37.5) | 29.7 |  |
| 60 | None | Half | Elementary teacher | 17.4 (3.4, 29.4) | 17.7 |  |
| 60 | None |  | Middle school teacher | 31 (12.5, 48.9) | 32.6 |  |
| 60 | None |  | High school teacher | 26 (5.4, 44.2) | 27.8 |  |
| 60 | Masks | Equal | Elementary student | 44.3 (0, 70) | 51 | 168 |
| 60 | Masks | Half | Elementary student | 12.2 (-0.3, 25.8) | 10 | 46 |
| 60 | Masks |  | Middle school student | 20.7 (-0.9, 41.5) | 19.5 | 87 |
| 60 | Masks |  | High school student | 4.7 (-0.5, 10.5) | 3.7 | 32 |
| 60 | Masks | Equal | Elementary teacher | 10.6 (0, 19.7) | 11.1 |  |
| 60 | Masks | Half | Elementary teacher | 3.8 (-0.9, 9.7) | 3.4 |  |
| 60 | Masks |  | Middle school teacher | 7.7 (-2.3, 17.8) | 6.7 |  |
| 60 | Masks |  | High school teacher | 2.6 (-2.1, 8.9) | 1.9 |  |
| 60 | Masks + Testing | Equal | Elementary student | 25.3 (-1, 55.9) | 22.5 | 96 |
| 60 | Masks + Testing | Half | Elementary student | 4.4 (-1, 12.1) | 2.3 | 17 |
| 60 | Masks + Testing |  | Middle school student | 8.5 (-1.5, 25) | 3.7 | 36 |
| 60 | Masks + Testing |  | High school student | 1.8 (-1, 5) | 1 | 12 |
| 60 | Masks + Testing | Equal | Elementary teacher | 5.6 (-0.9, 14.7) | 4.3 |  |
| 60 | Masks + Testing | Half | Elementary teacher | 1.2 (-1.8, 4.5) | 0.9 |  |
| 60 | Masks + Testing |  | Middle school teacher | 2.9 (-2.3, 11.1) | 2.2 |  |
| 60 | Masks + Testing |  | High school teacher | 0.9 (-3.9, 4) | 0 |  |
| 60 | Masks + Cohorts | Equal | Elementary student | 9.5 (-0.4, 20.1) | 7.9 | 36 |
| 60 | Masks + Cohorts | Half | Elementary student | 2.2 (-0.8, 5.1) | 1.7 | 8 |
| 60 | Masks + Cohorts |  | Middle school student | 2.9 (-1.2, 7.7) | 1.7 | 12 |
| 60 | Masks + Cohorts |  | High school student | 0.7 (-0.7, 2.2) | 0.5 | 5 |
| 60 | Masks + Cohorts | Equal | Elementary teacher | 1.6 (-0.9, 5.3) | 1.7 |  |
| 60 | Masks + Cohorts | Half | Elementary teacher | 0.5 (-1.8, 2.7) | 0.8 |  |
| 60 | Masks + Cohorts |  | Middle school teacher | 1 (-2.4, 4.5) | 0 |  |
| 60 | Masks + Cohorts |  | High school teacher | 0.3 (-3.8, 3.8) | 0 |  |
| 70 | None | Equal | Elementary student | 73.1 (44.8, 92.4) | 81.3 | 278 |
| 70 | None | Half | Elementary student | 31.2 (-0.2, 53.6) | 32.8 | 119 |
| 70 | None |  | Middle school student | 41.6 (9.4, 60.2) | 49.2 | 175 |
| 70 | None |  | High school student | 19 (-0.5, 32.3) | 20.9 | 129 |
| 70 | None | Equal | Elementary teacher | 22.5 (9.4, 34.5) | 24.1 |  |
| 70 | None | Half | Elementary teacher | 12.1 (0, 21.7) | 12.4 |  |
| 70 | None |  | Middle school teacher | 21.6 (0, 35.6) | 23.8 |  |
| 70 | None |  | High school teacher | 15.1 (0, 30.2) | 15.2 |  |
| 70 | Masks | Equal | Elementary student | 45 (9.7, 75.9) | 50.2 | 171 |
| 70 | Masks | Half | Elementary student | 9.6 (-0.5, 21.9) | 7.1 | 36 |
| 70 | Masks |  | Middle school student | 14.4 (-0.9, 34.1) | 10.6 | 61 |
| 70 | Masks |  | High school student | 1.8 (-0.5, 4.7) | 1.2 | 12 |
| 70 | Masks | Equal | Elementary teacher | 8.9 (0, 16.8) | 8.8 |  |
| 70 | Masks | Half | Elementary teacher | 2.5 (-1.8, 6) | 1.8 |  |
| 70 | Masks |  | Middle school teacher | 4.5 (-2.4, 13) | 2.3 |  |
| 70 | Masks |  | High school teacher | 1 (-2, 5.5) | 0 |  |
| 70 | Masks + Testing | Equal | Elementary student | 22.3 (-1, 54.5) | 16.5 | 85 |
| 70 | Masks + Testing | Half | Elementary student | 3.2 (-0.6, 9.6) | 1.1 | 12 |
| 70 | Masks + Testing |  | Middle school student | 5.1 (-1.1, 16.9) | 1.3 | 22 |
| 70 | Masks + Testing |  | High school student | 0.7 (-0.6, 2.2) | 0.3 | 5 |
| 70 | Masks + Testing | Equal | Elementary teacher | 4.2 (-1.8, 11.2) | 2.6 |  |
| 70 | Masks + Testing | Half | Elementary teacher | 0.8 (-1.8, 3.5) | 0.8 |  |
| 70 | Masks + Testing |  | Middle school teacher | 1.6 (-2.4, 6.7) | 0 |  |
| 70 | Masks + Testing |  | High school teacher | 0.3 (-2, 3.8) | 0 |  |
| 70 | Masks + Cohorts | Equal | Elementary student | 8.5 (-0.8, 19.3) | 6.3 | 32 |
| 70 | Masks + Cohorts | Half | Elementary student | 1.7 (-0.6, 4.3) | 1.1 | 6 |
| 70 | Masks + Cohorts |  | Middle school student | 2.1 (-1.2, 5.9) | 1 | 9 |
| 70 | Masks + Cohorts |  | High school student | 0.3 (-0.5, 1.2) | 0.2 | 2 |
| 70 | Masks + Cohorts | Equal | Elementary teacher | 1.2 (-0.9, 4.3) | 0.9 |  |
| 70 | Masks + Cohorts | Half | Elementary teacher | 0.3 (-1.8, 2.6) | 0 |  |
| 70 | Masks + Cohorts |  | Middle school teacher | 0.5 (-2.3, 4.4) | 0 |  |
| 70 | Masks + Cohorts |  | High school teacher | 0.2 (-2, 3.6) | 0 |  |

\*Assuming a 380-person elementary school, 420-person middle school, and 680-person high school

### Transmission model details

We developed a discrete-time, age-structured individual-based stochastic model to simulate COVID-19 transmission dynamics in the synthetic population (Figure 1A). At each point in time, representative of one day, each individual is associated with an epidemiological state: successfully vaccinated (V), susceptible (S), exposed (E), asymptomatic (A), symptomatic with non-severe illness (C), symptomatic with severe illness (H1, D1) resulting in eventual hospitalization before recovery (H2) or hospitalization before death (D2), recovered (R), or dead (M). Model parameters are in Table S1.

The daily contact rate between individuals *i* and *j* on day *t*, *K*_ij,t_, was estimated for pairs of individuals,

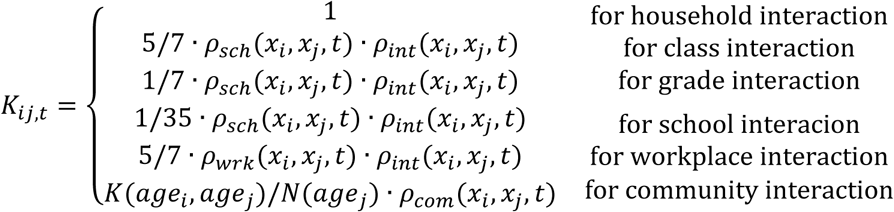

where the scaling ratios between classes, grades, and schools were obtained from previous study on transmission in various settings.[15] Community interaction represents the number of contacts expected between individuals from age groups of individuals *i* and *j* scaled by the number of individuals in the age group of individual *j*. *ρ*_*int*_(*x_i_, x_j_, t*) is a factor between 0 and 1 representing a social distancing intervention to reduce contact between individual pairs, and is equal to one under a no-intervention scenario. Because symptomatic individuals mix less with the community[16], we simulated a 100% reduction in daily school or work contacts and a 75% reduction in community contacts for a proportion (48%) of symptomatic individuals, and an additional proportion (50%) of their household members.[17] For these individuals, *ρ*_*sch*_(*x_i_, x_j_, t*) and *ρ*_*wrk*_(*x_i_, x_j_, t*) is equal to 0 and *ρ*_*com*_(*x_i_, x_j_, t*) is equal to 0.25, if: 1) either individual *i* or *j* is symptomatic (C, H1, or D1) on day *t* and isolates with some probability, or 2) either individual *i* or *j* is a household member of a symptomatic individual on day *t* and quarantines with some probability; and otherwise equal to 1. We assumed that individuals were in the infectious class for up to 3 days prior to observing symptoms[18], during which time they did not reduce their daily contacts.

Transmission was implemented probabilistically for contacts between susceptible (S) and infectious individuals in the asymptomatic (A) or symptomatic and non-hospitalized states (C, H1, D1). Movement of individual *i* on day *t* from a susceptible to exposed class is determined by a Bernoulli random draw with probability of infection per day given by the daily force of infection, λ_i,t_:

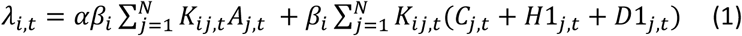

where *α* is the ratio of the force of infection between asymptomatic and symptomatic individuals; and *β*_*i*_is calculated from 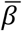, the population mean transmission rate of the pathogen. 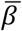 is determined using the next-generation matrix method[19] as:

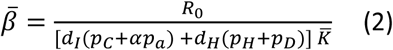

where *R*_0_ is the basic reproduction number (defined as the expected number of secondary cases from a single infected case in a completely susceptible population); p_s_ is the proportion of agents destined for state *s*; d_I_ is the average time between infection and recovery for tracks A and C; d_H_ is the average time between infection and hospitalization for tracks H and D; and 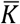 is the mean number of contacts an individual makes daily under no interventions, weighted by their probability of being contacted.[20] Here, we calculated *R*_0_ as 4.6, based on an average of *R*_0_ for the Alpha (*R*_0_ = 2.5 and proportion = 16%) and Delta variant (*R*_0_ = 5.0 and proportion = 84%), weighted by the proportion of circulating variants in summer 2021 [1, 2]. We represent age-varying susceptibility[9] using an age-stratified *β_i_* that incorporates the ratio of the susceptibility of adults to children and jointly solves equations (3) and (4):

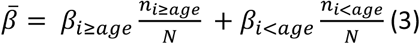

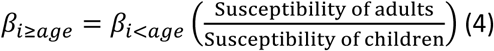

The duration of the latent period, d_L_, for each individual transitioning from class E was drawn from a Weibull distribution with mean 5.4 days (95% CI: 2.4, 8.3).[3–5] Whether an individual remained asymptomatic, or was hospitalized, or died was determined via Bernoulli random draws from age-stratified conditional probabilities (Figure 1B, Table S5). The time to recovery for non-hospitalized cases (mean: 13.1 days, 95% CI: 8.3, 16.9)[6], the time to hospitalization for severe cases (mean: 10.3, 95% CI: 6.5, 13.3)[7], and time to recovery or death for hospitalized cases (mean: 14.4, 95% CI: 11.3, 16.6) were sampled from Weibull distributions (Table S5).[8]

### Description of reopening strategies

#### 1. Schools open without precautions

In this scenario, schools are open under a business-as-usual scenario. For all interactions, *ρ*_*int*_(*x_i_, x_j_, t*) = 1. The average class size is 20 students, the average sizes of elementary (K - 5), middle (6-8), and high schools (9-12) are 380, 420, and 620 students.

#### 2. Students and faculty wear masks

In this scenario, we assume that both students and teachers wear masks while at school. We assume that the masks both reduce the likelihood of acquiring COVID-19, as well as the likelihood of transmitting it. We assume that the effectiveness of masks for elementary school children is 15%, the effectiveness for middle school children is 25%, the effectiveness for high school children is 35% and the effectiveness for teachers is 50%. Accordingly, for each school, grade, or class pair, we have:

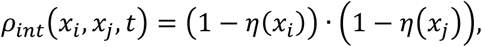

where *η*(*x*_*i*_) represents the effectiveness of the mask for individual *i.* such that *η*(*x*_*i*_) = 0.15 if the individual is an elementary school student, *η*(*x*_*i*_) = 0.25 if the individual is a middle school student, *η*(*x*_*i*_) = 0.35 if the individual is a high school student, and *η*(*x*_*i*_) = 0.5 if the individual is a teacher or staff member.

#### 3. <u>Stable cohorts:</u> *classroom groups are enforced, reducing other grade and school contacts by 75%*

In this scenario, we assume that students reduce their contacts with other teachers and students outside of their class group (or cohort) by a given proportion. We model both reductions of outside-class contacts by 50% (“weak” cohort approach) or 75% (“strong” cohort approach). The size of the class group is 20 students, on average. This may be equivalent to reductions in lunchroom or recess contacts, while still permitting chance interactions in the hallways or bathrooms. Here, we update *ρ*_*int*_(*x_i_, x_j_, t*) such that:

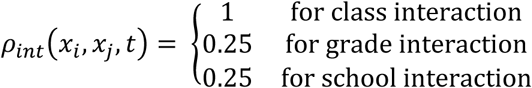

#### 4. <u>Weekly testing of teachers and students (periodic test-trace-isolate, TTI):</u> *Faculty and students are tested with 85% sensitivity on a weekly or monthly basis^42^, and positive cases are isolated and their class quarantined for 14 days*

In this scenario, every 7 or 30 days, the state of the non-hospitalized agents are ascertained through a simulated test. We assumed that the test would detect individuals in a symptomatic or asymptomatic or pre-symptomatic state with 85% sensitivity and 100% specificity. If a truly positive case was simulated to test positive, the case would reduce their school contacts by 100% for 14 days and their community contacts by 75% for 14 days. Additionally, the students or teacher in the same class as the case would reduce their school contacts by 100% and their community contacts by 75% for 14 days. This is implemented though updating *ρ*_*sch*_(*x_i_, x_j_, t*) and *ρ*_*comm*_(*x_i_, x_j_, t*) as described. If a school administrator tested positive, only the administrator isolated for 14 days

### Supplemental results for Alpha variant

#### Effect of within-school precautions under various community vaccination coverages – Alpha variant

**Table S8.** Excess symptomatic infections attributable to school transmission assuming circulation of the Alpha variant only. Results are stratified by levels of community vaccination coverage and within-school non-pharmaceutical intervention (NPI).

| NPI | Susceptibility of children <10 years | Population | 50% coverage |  | 60% coverage |  | 70% coverage |  |
| --- | --- | --- | --- | --- | --- | --- | --- | --- |
|  |  |  | Mean (89% HPDI) excess infections per 100 population | Median excess infections per 100 population | Mean (89% HPDI) excess infections per 100 population | Median excess infections per 100 population | Mean (89% HPDI) excess infections per 100 population | Median excess infections per 100 population |
| None | Equal | Elementary student | 2.4 (-0.2, 6.9) | 1.4 | 1.8 (-0.2, 5.5) | 0.7 | 1.3 (-0.2, 4.3) | 0.2 |
| None | Half | Elementary student | 0.5 (-0.2, 1.2) | 0.2 | 0.3 (-0.1, 1) | 0.1 | 0.2 (-0.1, 0.7) | 0 |
| None |  | Middle school student | 2.4 (-0.2, 7.9) | 0.6 | 1.4 (-0.2, 5.4) | 0 | 0.8 (-0.2, 3) | 0 |
| None |  | High school student | 1 (-0.2, 3.6) | 0.2 | 0.3 (-0.2, 1) | 0 | 0.1 (0, 0.3) | 0 |
| None | Equal | Elementary teacher | 2.8 (-0.9, 8.4) | 1.7 | 1.8 (-0.9, 6.7) | 0.9 | 1 (-0.9, 4.3) | 0 |
| None | Half | Elementary teacher | 0.8 (-1.8, 2.7) | 0.8 | 0.4 (-0.9, 2.6) | 0 | 0.2 (-0.9, 1.7) | 0 |
| None |  | Middle school teacher | 4.1 (-2.4, 14.9) | 0 | 2 (-2.3, 8.9) | 0 | 0.9 (-2.3, 4.4) | 0 |
| None |  | High school teacher | 2.4 (-2, 9.4) | 0 | 0.7 (-2, 3.8) | 0 | 0.2 (-1.9, 1.9) | 0 |
| Masks | Equal | Elementary student | 0.5 (-0.2, 1.6) | 0.2 | 0.4 (-0.2, 1.1) | 0.1 | 0.3 (-0.2, 0.9) | 0 |
| Masks | Half | Elementary student | 0.1 (-0.1, 0.3) | 0 | 0.1 (-0.1, 0.3) | 0 | 0.1 (-0.1, 0.2) | 0 |
| Masks |  | Middle school student | 0.2 (-0.3, 0.7) | 0 | 0.1 (-0.2, 0.4) | 0 | 0.1 (0, 0.4) | 0 |
| Masks |  | High school student | 0.1 (-0.2, 0.3) | 0 | 0 (0, 0.3) | 0 | 0 (0, 0.2) | 0 |
| Masks | Equal | Elementary teacher | 0.4 (-0.9, 2.5) | 0 | 0.2 (-0.9, 1.8) | 0 | 0.1 (-0.9, 1.7) | 0 |
| Masks | Half | Elementary teacher | 0.1 (-0.9, 1.8) | 0 | 0.1 (-0.9, 1.7) | 0 | 0 (-0.9, 0.9) | 0 |
| Masks |  | Middle school teacher | 0.3 (-2.3, 2.3) | 0 | 0 (-2.3, 2.2) | 0 | 0.1 (-2.3, 2.2) | 0 |
| Masks |  | High school teacher | 0.1 (-2, 1.9) | 0 | 0 (-1.9, 1.9) | 0 | 0 (-1.9, 1.9) | 0 |

**Figure S1.**
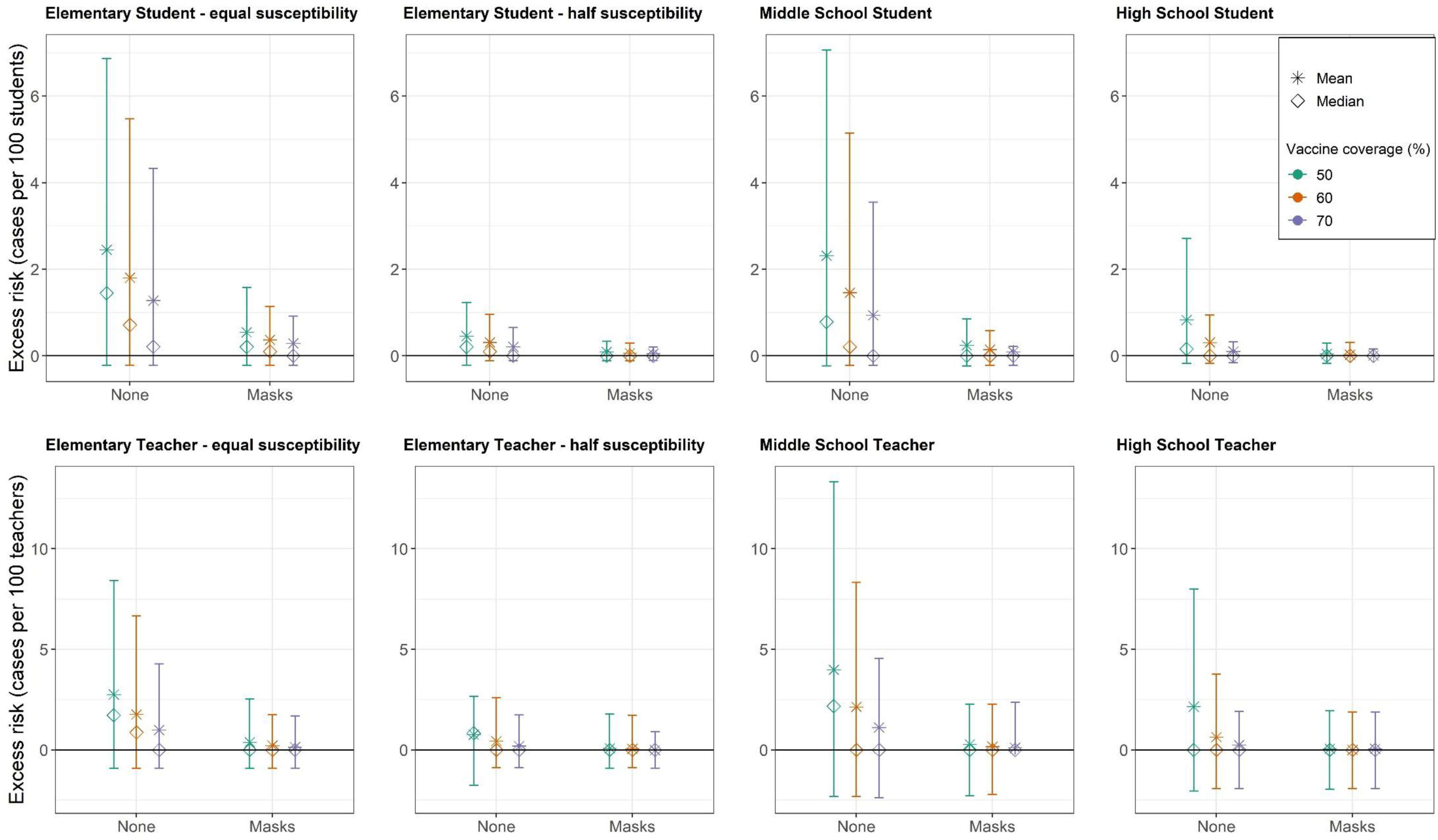
We examined the effect of masks across three levels of community vaccination coverage (50%, 60%, 70%), assuming circulation of the Alpha variant only, and that the vaccine effectiveness is 85% against symptomatic infection. We examined the results across two assumptions about the susceptibility of children (children <10 half as susceptible to SARS-CoV-2 as those 10+ vs. equally susceptible across all ages). We calculated the mean (stars) and median (diamonds) of excess cases per 100 persons attributable to school transmission among population subgroups across 1,000 model realizations. Vertical lines reflect the 89^th^percentile high probability density interval (HPDI).

#### Interventions required to reduce incidence attributable within schools below certain risk thresholds – Alpha variant

**Table S9.** The minimum non-pharmaceutical intervention(s), or minimum within-school vaccination coverage of the eligible population, needed to reduce the risk of symptomatic infection to beneath a given threshold (e.g., 50 cases per 1,000 population), assuming that 70% of the vaccine-eligible community has received a vaccine at 85% effectiveness. Simulations examine circulation of the Alpha variant alone.

|  |  | Threshold - symptomatic cases per 1,000 population |  |  |  | < 1 case per school** |
| --- | --- | --- | --- | --- | --- | --- |
|  |  | <50 | <10 | <5 | <1 |  |
| Students | Elementary school – equal susceptibility | 70% within-school coverage | Masks | Masks | Masks + cohorts | Masks |
|  | Elementary school – half susceptibility | 70% within-school coverage | 70% within-school coverage | Masks or 85% within-school coverage | Masks | Masks |
|  | Middle school | 70% within-school coverage | 70% within-school coverage | Masks or 95% within-school coverage | Masks | Masks |
|  | High school | 70% within-school coverage | 70% within-school coverage | 70% within-school coverage | Masks or 75% within-school coverage | 70% within-school coverage |
| Teachers | Elementary school – equal susceptibility | 70% within-school coverage | 70% within-school coverage | Masks or 90% within-school coverage | Masks |  |
|  | Elementary school – half susceptibility | 70% within-school coverage | 70% within-school coverage | 70% within-school coverage | Masks or 85% within-school coverage |  |
|  | Middle school | 70% within-school coverage | 70% within-school coverage | Masks or 90% VC | Masks |  |
|  | High school | 70% within-school coverage | 70% within-school coverage | 70% within-school coverage | Masks or 85% within-school coverage |  |

\*not observed under the specific combination of interventions simulated
\*\*Assuming a 380-person elementary school, 420-person middle school, and 680-person high school

